# Hypothalamic structural differences link sleep and cognition across the Alzheimer’s disease spectrum

**DOI:** 10.64898/2026.03.11.26346020

**Authors:** Weiran Zhang, Nicole Espinosa, Aaron Lam, Angela L D’Rozario, Sharon Naismith, Nathan Cross

## Abstract

**INTRODUCTION:** Sleep disturbance is common across the Alzheimer’s disease (AD) spectrum, yet the structural substrates linking sleep disruption and cognitive decline remain unclear. The hypothalamus regulates sleep–wake function and is affected by AD pathology, but its role across clinical stages is poorly defined.

**METHODS:** 672 older adults classified as subjective cognitive decline, single- and multi-domain mild cognitive impairment, or probable AD underwent structural MRI. Subjective insomnia measures were available for the full cohort, and 209 underwent polysomnography. Associations between hypothalamic volume, sleep architecture, and cognition were examined.

**RESULTS:** Hypothalamic volume declined progressively from SCD to MD-MCI and AD, with the greatest reductions in anterior subregions. Smaller hypothalamic volume associated with diminished slow-wave sleep, lower REM sleep in SD-MCI, poorer neuropsychological functions, and moderated the association between hippocampal volume and memory.

**CONCLUSION:** Hypothalamic atrophy emerges along the AD continuum, and relates to specific alterations in sleep quality and cognition.

## Background

Alzheimer’s disease (AD), the leading cause of dementia in older adults, is conceptualised as a clinical continuum spanning subjective cognitive decline (SCD), mild cognitive impairment (MCI), and dementia. MCI may present with memory impairment (amnestic MCI) and may involve single (SD-MCI) or multiple (MD-MCI) cognitive domains, with the amnestic and multiple-domain types conferring a higher risk of progression to AD [1,2].

Sleep disturbances and disorders are quite common (up to 66%) among patients with AD, relative to older adults generally [3–6]. Individuals with MCI and AD exhibit alterations in sleep architecture, including reduced slow-wave sleep (SWS) and rapid eye movement (REM) sleep [7], as well as worse severity of hypoxemia relative to healthy older adults [8,9]. However, it remains unclear whether such changes primarily reflect downstream effects of neurodegeneration or disruption of brain regions that regulate sleep–wake function.

The hypothalamus is a small nucleus of the brain, playing a central role in in circadian regulation, arousal, sleep initiation and maintenance, as well as endocrine regulation, appetite and stress response [10,11]. This is attributed to the multiple sub-nuclei that comprise the hypothalamus, each with varying functions. Sleep-promoting nuclei, including the ventrolateral preoptic nucleus (VLPO), interact with the wake-promoting systems such as orexinergic neurons in the lateral hypothalamus, and also with the regulator of the circadian timing system - suprachiasmatic nucleus (SCN) to maintain sleep–wake balance and the 24-hour circadian rhythm [12–15]. Previous studies have confirmed that the activation of VLPO galanin neurons promotes NREM sleep [16], and that damage to the VLPO leads to insomnia [17]. The orexin system in the lateral hypothalamus, composed of orexinergic cells, plays a key role in promoting wakefulness and a reduction in orexinergic cells can result in narcolepsy [18].

Neuropathological and neuroimaging studies indicate that the hypothalamus is impacted by AD, with evidence of tau and amyloid pathology [19], and structural volume reductions in MCI and dementia [20–22]. It has been proposed that the loss of integrity and neural dysfunction in the SCN may be a result of, or even a contributing factor to, the development of AD [23]. However, the clinical stage at which hypothalamic structural differences become detectable, and their functional relevance for sleep disturbance and cognition, remain poorly defined.

An important unresolved question is whether hypothalamic changes reflect generalised subcortical neurodegeneration or a region-specific vulnerability that could contribute to the myriad of sleep disturbances in AD. The hippocampus and thalamus undergo early structural changes in AD and provide benchmarks against which to assess the specificity of hypothalamic effects [24–26]. The hypothalamus also plays a critical role in cognition via integration within hippocampal-limbic circuits and regulation of the hypothalamic-pituitary-adrenal axis, which has been linked to age-related cognitive decline [27,28]. Thus, it is possible that the hypothalamus may moderate the relationship between hippocampal atrophy and memory decline seen in AD.

The objective of this study was to observe whether structural hypothalamic volumes differ across the clinical stages of SCD, SD-MCI, MD-MCI and probable AD. Given its role in regulating sleep and wakefulness, we also aimed to investigate the associations between hypothalamic volume and measures of sleep quality and architecture (e.g. sleep efficiency, sleep latency, REM latency, REM and non-REM sleep). We hypothesised that hypothalamic volume is inversely proportional to the severity of cognitive impairment, being smallest in AD and following similar patterns to hippocampal and thalamic atrophy. Secondly, hypothalamic volume (especially the anterior and lateral sub-regions) would predict both objective and subjective sleep quality and be stronger predictors than hippocampal or thalamic volume. Third, hypothalamic volume may support memory by mediating the link between hippocampal volume and memory across the neurodegenerative spectrum.

## Methods

Participants were recruited from the Healthy Brain Ageing Clinic at the Brain and Mind Centre in the University of Sydney, Australia. This is a specialist assessment and intervention clinic for older people with cognitive and/or mood concerns. Inclusion criteria were age ≥ 50 years, referral for cognitive assessment, and any participant who had been referred for a sleep study. Exclusion criteria were any neurological disease (e.g. Parkinson’s disease, epilepsy), a history of (or current) psychosis, prior stroke or head injury (with loss of consciousness > 30 minutes), alcohol or substance misuse (> 14 standard drinks), sleep disorders (e.g. narcolepsy) or shiftwork, or inadequate English to complete neuropsychological testing. Approval for this study was granted by the University of Sydney Human Research Ethics Committee, and all participants gave written informed consent prior to study participation. This observational study is reported in accordance with the STROBE (Strengthening the Reporting of Observational Studies in Epidemiology) guidelines.

### Clinical Assessment

#### Medical assessment

A medical specialist recorded medical and sleep disorders history, alcohol (units per week) and medication use, and completed anthropometric data for computing body mass index (BMI). Medical burden was rated using the Cumulative Illness Rating Scale – Geriatric Version (CIRS-G) [29], higher CIRS-G score refers to medical burden. Depressive symptoms were reported using the 15-item Geriatric Depression Scale (GDS-15) [30], where higher scores indicate greater depressive symptoms (range: 0 to 15).

#### Neuropsychological assessment

As detailed elsewhere [31], a clinical neuropsychologist conducted a standardised neuropsychological assessment for each participant. Briefly, this battery included the Wechsler Test of Adult Reading (WTAR) [32], Trail Making Test, Part A and B (TMT-A and TMT-B) [33], Wechsler Adult Intelligence Scale - Third Edition (WMS-III) Logical Memory (% retention) [34], Boston Naming Test (BNT) [35], and the Controlled Oral Word Association Test (FAS; COWAT) [36]. Rey Auditory Verbal Learning Test (RAVLT) was used to assess verbal declarative memory performance [37]. From this test, we defined encoding ability as the summed score for the five learning trials (a1–5), and delayed recall as the number of words recalled at delay (a7). All scores were converted to z-scores or scaled scores using age and (where relevant) education adjusted normative.

For the primary and exploratory aspects of the paper, as previously described [38], we computed age and (where relevant) education adjusted z-scores and then derived composite scores for:

- Verbal memory = (Rey Auditory Verbal Learning Test [RAVLT] trial A7 + Wechsler Memory Scale III Logical Memory II)/2
- Executive functioning = ([DKEFS Color Interference Word Test 3 + 4]/2 + Controlled Oral Word Association Test (letters F,A,S) + Trail Making Test Part B)/3
- Processing Speed = ([DKEFS Color Interference Word Test 1+2]/2 + Trail Making Test Part A)/2

#### MCI and SCD classifications

Individuals were classified as having clinical MCI according to Winblad’s criteria [39], with classifications and MCI subtypes consensus rated by a panel consisting of two clinical neuropsychologists and a medical specialist (Geriatrician or Neurologist). MCI patients with only one impaired symptom listed above (e.g. language impairment only) were classified as single-domain MCI, while MCI patients with at least two impaired functions were classified as multi-domain MCI. Additionally, MCI patients were classified as amnestic (aMCI) or non-amnestic (naMCI), based on the presence of memory deficits. Those who did not exhibit neuropsychological evidence of cognitive decline or major depression were included as SCD group [40]. A probable diagnosis of AD was made by the clinical team based off neuropsychological testing and clinical assessment by the medical specialist [41,42].

### Brain structure

#### MRI acquisition parameters

Participants underwent T1-weighted magnetic resonance imaging (MRI). Scans were conducted within six-months of clinical assessment (mean 26 days) and took place at two different facilities in Sydney, Australia (for more details on scanning parameters see supplementary materials).

#### Segmentation of brain regions

Volumetric segmentation and labelling were automatically estimated by the FastSurfer toolbox using differences in voxel intensity and registration to a probabilistic atlas [43]. The automated segmentation tool in FreeSurfer 7.4.1, which employs a validated deep-learning-based approach [22], was employed to segment the hypothalamus and its five subunits: (1) anterior–inferior hypothalamus, which contains preoptic area and paraventricular nucleus (PVN); (2) the anterior–superior hypothalamus, which contains SCN and supraoptic nucleus (SON); (3) the posterior hypothalamus, which contains mamillary body (including medial and lateral mamillary nuclei), lateral hypothalamus, tuberomamillary nucleus (TMN); (4) the inferior tubular hypothalamus, which contains infundibular (or arcuate) nucleus, ventromedial nucleus, SON, lateral tubular nucleus and TMN; and (5) the superior tubular hypothalamus, which contains dorsomedial nucleus, PVN and lateral hypothalamus (Figure 1). The segmentation maps were visually examined to verify their accuracy and precision. Each participant’s estimated total intracranial volume (eTIV) was also collected for correction and normalisation of hypothalamus volume. To compare neurodegenerative rates of other brain regions across groups, bilateral volumetric measures (mm^3^) of the hippocampus and thalamus were investigated using the automatic subcortical segmentation. For more details see Espinosa et al. [38].

**Figure 1.**
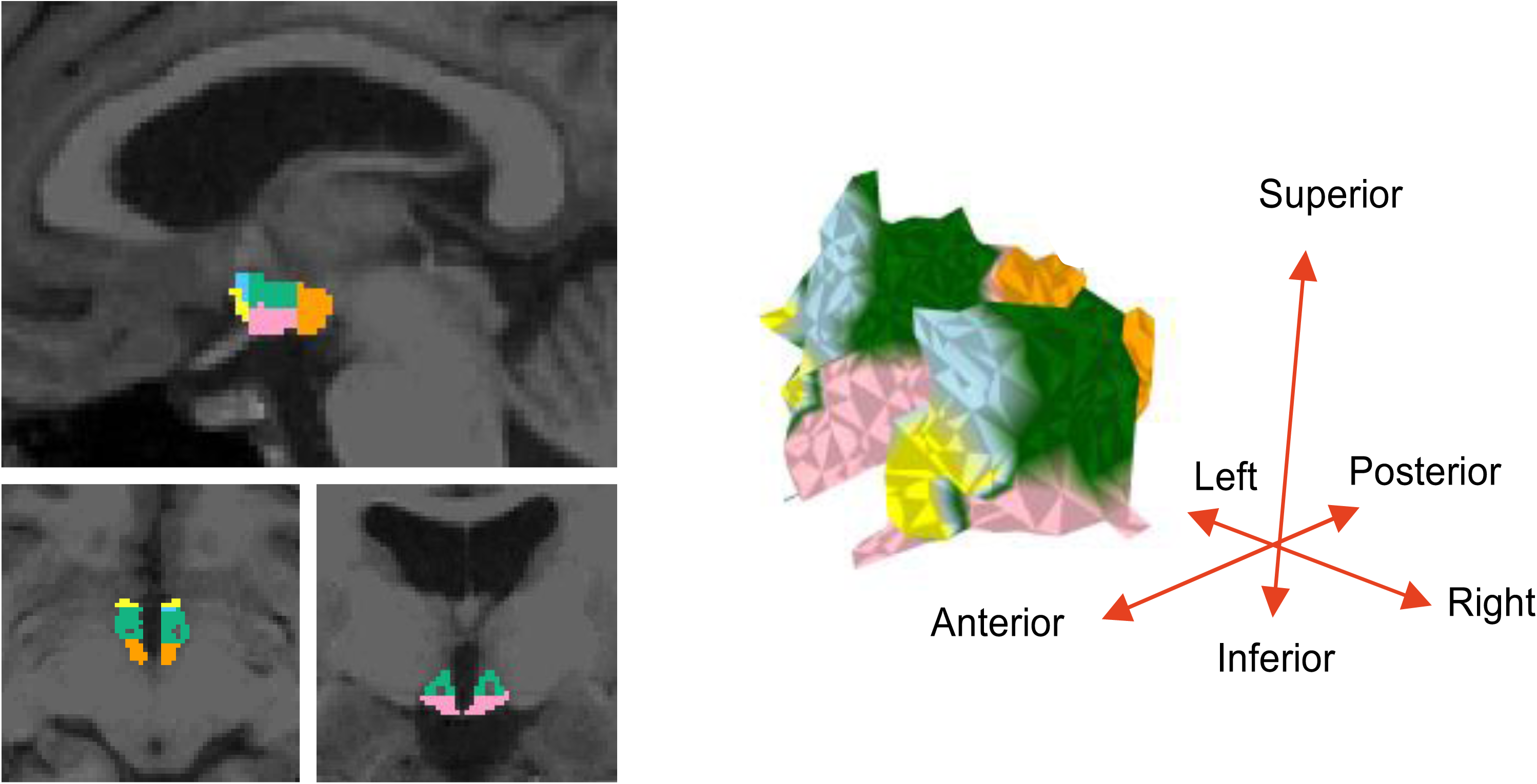
Left: Segmentation of the hypothalamus on structural MRI. Right: The five subregions of the hypothalamus: anterior superior (blue), anterior inferior (yellow), superior tubular (green), inferior tubular (pink) and posterior (orange).

### Sleep quality

#### Subjective sleep quality

Self-perceived sleep quality was assessed using the Pittsburgh Sleep Quality Index (PSQI) [44]. In addition to the PSQI global score, this self-report tool can be broken down into subcomponents to capture specific dimensions of subjective sleep quality. Although the global score provides an overall index of sleep disturbance, it may obscure differential relationships between distinct aspects of sleep (e.g., latency, duration, sleep disturbances) and neurobiological measures. Recent work demonstrates that relying solely on the global PSQI score can mask meaningful variation across sleep dimensions and limit insight into sleep–brain associations [45]. The PSQI subcomponents are:

*Component 1* - subjective sleep quality

*Component 2* - sleep latency (time taken to fall asleep)

*Component 3* - sleep duration (total hours of sleep per night)

*Component 4* - sleep efficiency (percentage of time in bed spent sleeping)

*Component 5* - sleep disturbances (frequency of awakenings or discomfort)

*Component 6* - use of sleep medication (frequency of pharmacological interventions)

*Component 7* - daytime dysfunction (effect of sleep problems on daytime functions)

Each component is rated on a scale from 0 to 3, with higher scores indicating greater impairment. The sum of the seven components yields a global PSQI score ranging from 0 to 21, where scores above 5 typically indicate poor sleep quality.

#### Overnight polysomnography

A subset (n = 209) of participants underwent a full, overnight, in-laboratory sleep study (Embla Titanium or Sandman, Natus, CA, USA; Alice-5, Philips Respironics, Murrysville, PA, USA; or Grael, Compumedics Siesta, Abbotsford, VIC, Australia) at the Woolcock Institute of Medical Research within 6 months of the neuropsychological assessment and MRI scan (mean 8.3 days). Sleep studies used either a standard or research (6 or 10 channel EEG) PSG montage with electrooculogram, chin and leg electromyogram, electrocardiogram, nasal airflow pressure (nasal cannula), thoracic and abdominal respiratory effort, finger pulse oximetry (SPO_2_) and body position. For more details, see Lam et al. [46].

Sleep staging and scoring were performed using AASM V2.3 standardised criteria by registered sleep technologists [47]. Sleep architecture measures including total sleep time (TST), sleep efficiency, sleep latency, rapid eye movement (REM) latency, and duration of REM and NREM (N1, N2 and N3 sleep) sleep stages (% TST) were obtained. Obstructive sleep apnoea (OSA) data including apnoea-hypopnea index (AHI), EEG arousal index (ArI), mean oxygen saturation (SPO_2_) level and oxygen desaturation index 3% (ODI) were also collected.

### Statistical analysis

Data were analysed using R (Version 4.41). For group comparisons, chi-square method was applied for contingencies, ANOVA was used for parametric data, and Kruskal–Wallis was employed for non-parametric data [48]. Post-hoc pairwise analyses were conducted utilising either independent t-tests or the non-parametric Dunn test [49]. Spearman’s rank order coefficients were applied for non-parametric correlations. Corrections for multiple comparisons were conducted employing the Benjamini-Hochberg false discovery rate (FDR) procedure with the FDR threshold (q-value) set at 0.05 in a family-wise manner [50].

As age is known to influence brain structure, and preliminary analyses confirmed age differences across diagnostic groups (Table 1) as well as associations with hypothalamic volume (Supplementary Figure 1), age adjustment was applied prior to group and association analyses. Linear regression was used to remove age-related variance from all variables of interest, and the residuals were then used in subsequent analyses to examine the relationships among variables independent of age.

To examine regional differences in volumetric atrophy across diagnostic groups, we employed a linear mixed-effects model using the lmer function from the lme4 package in R. The dependent variable was z-scored regional brain volume, standardized using the mean and standard deviation of the SCD group to allow for cross-region comparison relative to a healthy baseline. The fixed effects included diagnostic group, region of interest (hippocampus, hypothalamus, thalamus), and their interaction. Random intercepts were modelled for each participant to account for within-subject correlations due to repeated measurements across regions. Model parameters were estimated using restricted maximum likelihood (REML), and p-values for fixed effects were calculated using Satterthwaite’s approximation for degrees of freedom, as implemented in the lmerTest package. Exploratory analysis were performed comparing participants in amnestic vs non-amnestic MCI to examine whether hypothalamic volume differences were consistent across alternative MCI subclassifications.

Principal components analysis (PCA) was used to reduce correlated PSG measures into orthogonal components that capture shared variance across tests. To assess whether the relationship between hypothalamus volume and objective sleep measures differed by diagnostic group, we fitted a linear model with an interaction term between volume and group (e.g. volume × group). This model tested for group differences in both the strength and direction of the association between regional brain volume and the outcome variable (e.g. PCA-derived sleep component).

To compare the strength of two dependent correlations (e.g. hypothalamus vs hippocampus) sharing one variable in common (e.g., verbal memory), we employed Steiger’s Z test [51], a modification of Dunn and Clark’s procedure that accounts for the dependency between correlations [52]. This test evaluates whether the difference between two correlation coefficients (e.g., hypothalamus–memory and hippocampus–memory) is statistically significant, using the average correlation and sample size to compute a z-statistic. All comparisons were conducted using the cocor package in R.

Finally, as three different types of MRI head coils (8, 20 and 32 channel) were employed to collect MRI data, linear models were fitted with an interaction term between volume and group and between PSG components and volume to assess the impact of head coil on the main findings.

## Results

A total of 672 participants were recruited, categorized into four clinical diagnostic groups: SCD (n = 226), SD-MCI (n = 127), MD-MCI (n = 266) and AD (n = 53). Demographics are shown in **Table 1**. The mean age across groups varied, as the AD group was significantly older (χ² = 46.5, df = 3, p < 0.001). The biological distribution also differed across the groups with the SCD group comprising more females. The education years in AD group were significantly fewer than the SCD (z = -2.8, p_FDR_ = 0.01) and MD-MCI groups (z = -3.1, p_FDR_ = 0.01). As expected, the MMSE scores demonstrated a decreasing trend from SCD and SD-MCI to MD-MCI to AD.

All cognitive function composites (verbal learning, verbal memory, executive function, language and processing speed) showed a decreased trend from SCD to SD-MCI to MD-MCI to AD.

### Subjective and objective sleep quality

The PSQI global score in AD group was significantly lower than the SCD, SD-MCI and MD-MCI groups (see Table 2). This effect was observed across all sub-domains of the PSQI, except for Component 7 (Daytime dysfunction).

For PSG-derived objective measure of sleep quality, the AD group showed a non-significant trend for reduced sleep efficiency (χ² =7.0, df = 3, p = 0.07) compared to both SD-MCI (Z = - 2.6, p = 0.01, p_FDR_ = 0.06) and MD-MCI (Z = -2.3, p = 0.02, p_FDR_ = 0.058).

#### Hypothalamus volume

Both the left and right hypothalamus volume differed across SCD, SD-MCI, MD-MCI and AD groups (left: χ² = 19.4, df = 3, p < 0.001Cright: χ² = 22.7, df = 3, p < 0.001, **Figure 2**). In the left hypothalamus, post-hoc pairwise comparisons using Dunn’s test, with a Benjamini-Hochberg correction for multiple comparisons, showed significant differences between AD and SCD (z = -4.0, p_FDR_ < 0.001), AD and SD-MCI (z = -3.5, p_FDR_ = 0.002), AD and MD-

**Figure 2.**
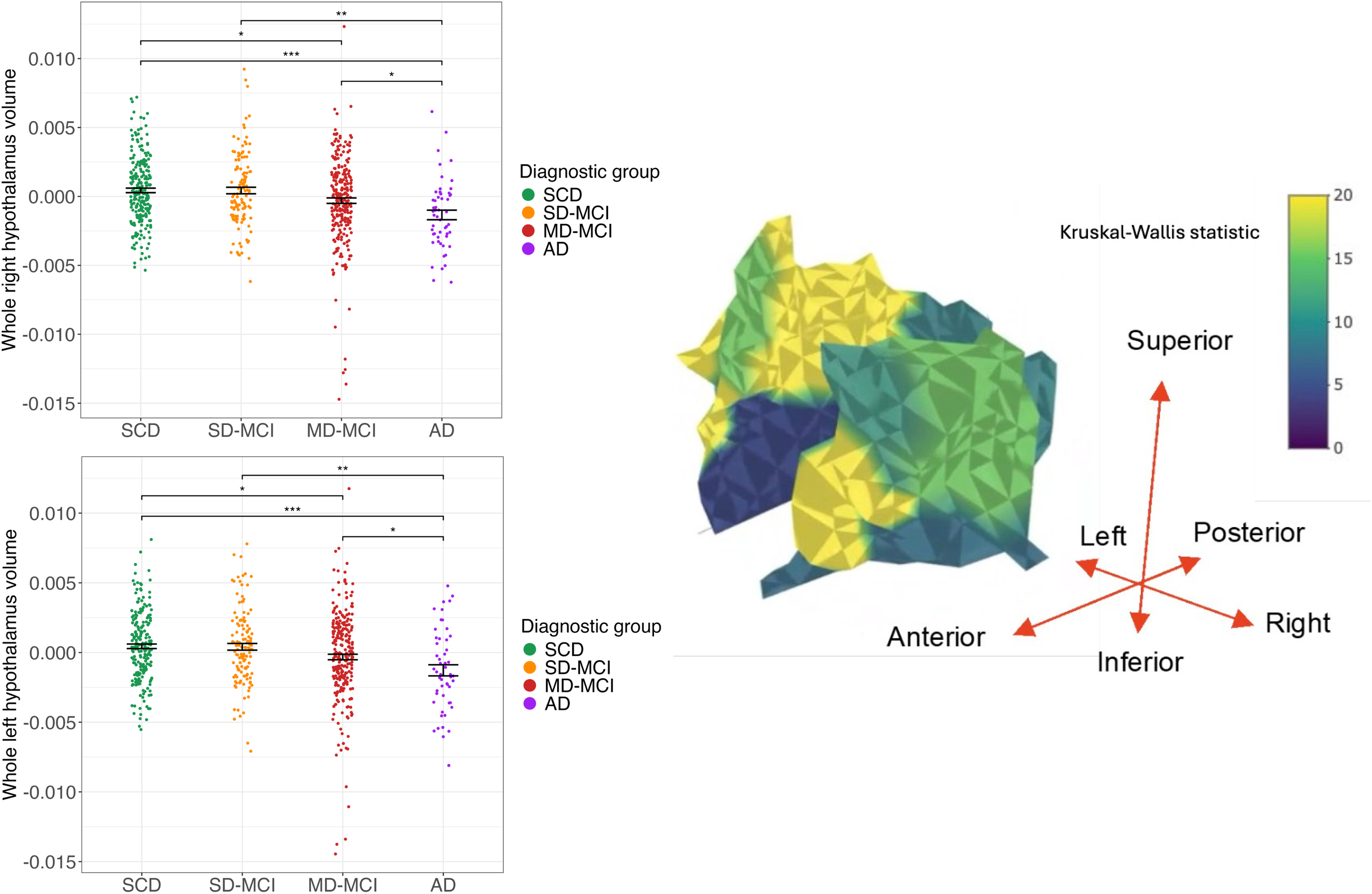
Hypothalamus volume in the left and right hemisphere across SCD, SD-MCI, MD-MCI and AD groups. Regional differences showed during the neurodegenerative process, anterior superior and tubular superior showed larger atrophy rate than posterior and tubular inferior.

MCI (z = -2.5, p_FDR_ = 0.02), MD-MCI and SCD (z = -2.5, p_FDR_ = 0.02). Similar differences were observed in the right hypothalamus between AD and SCD (z = -4.5, p_FDR_ < 0.001), AD and SD-MCI (z = -4.0, p_FDR_ < 0.001), and AD and MD-MCI group (z = -3.3, p_FDR_ = 0.002).

After controlling head coil, significant differences in hypothalamic volume were still observed across different diagnostic groups (See Supplementary Materials).

When MCI groups were reclassified into amnestic and non-amnestic subtypes, robust differences in hypothalamic volume were again observed, particularly between AD and SCD, and between aMCI and SCD/naMCI (see Supplementary Figure 2).

#### Regional hypothalamus volumes

Across all anterior, tubular, and posterior hypothalamic subregions, diagnostic groups differed significantly, with the smallest volumes consistently observed in the AD group and intermediate reductions in MD-MCI. These effects were most pronounced in the anterior hypothalamus, where AD showed robust bilateral reductions relative to all other groups, and MD-MCI also demonstrated smaller volumes than SCD and, in some cases, SD-MCI.

Subregional patterns were broadly consistent across hemispheres, and only the tubular inferior region showed weaker evidence for group differences. Full statistical results and pairwise contrasts are provided in the Supplementary materials (See Supplementary Figures 3-7).

#### Comparative atrophy rates in Hippocampus and Thalamus

Group differences in hippocampal volume revealed significant variation (χ² = 81.8, df = 3, p < 0.001, **Figure 3A**). The AD group had reduced hippocampal volumes compared to the MD-MCI (Z = -5.6, p_FDR_ < 0.001), SD-MCI (Z = -6.8, p_FDR_ < 0.001), and SCD (Z = -8.6, p_FDR_ < 0.001) groups. Furthermore, the MD-MCI group had smaller volumes than both the SD-MCI (Z = -2.4, p = 0.02) and SCD (Z = -5.1, p_FDR_ < 0.001) groups. There was a robust correlation between hypothalamus volume and hippocampus volume (rho = 0.58, p_FDR_ < 0.001, **Figure 3B**), mostly driven by anterior and tubular regions (see Supplementary Figure 8).

**Figure 3.**
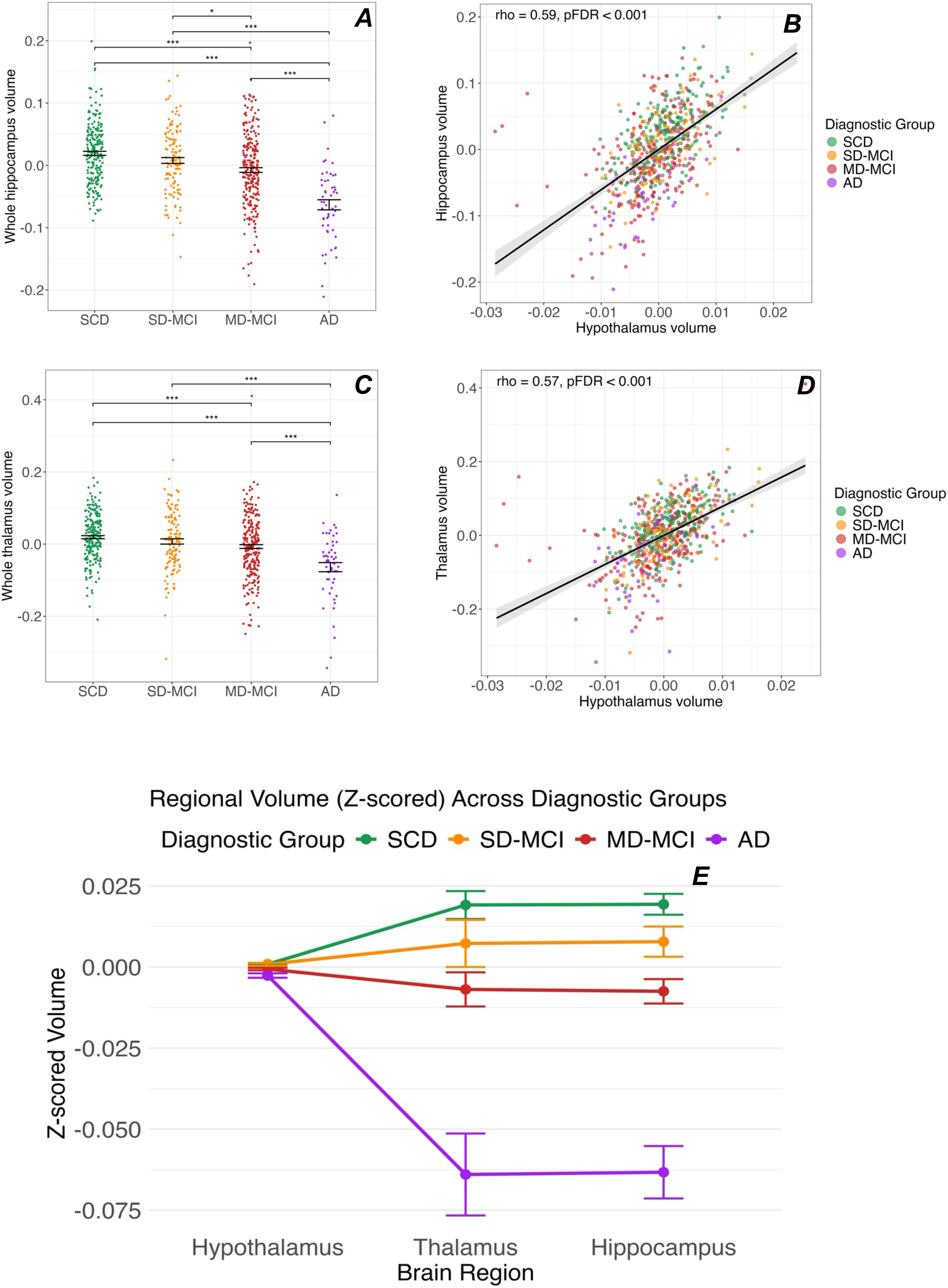
Relative differences of hippocampus, thalamus and hypothalamus across diagnostic groups. *Top Left (A)*: Hippocampus volume differences across SCD, SD-MCI, MD-MCI and AD groups. *Top Right (B)*: Hippocampus volume was associated with hypothalamus volume in the entire sample and within each diagnostic group. *Middle Left (C)*: Thalamus volume differences across SCD, SD-MCI, MD-MCI and AD groups. *Middle Right (D)*: Thalamus volume was associated with hypothalamus volume in the entire sample and within each diagnostic group. *Bottom (E)*: Mean z-scored volumes (± standard error) are plotted for the hippocampus, hypothalamus, and thalamus across diagnostic groups: subjective cognitive decline (SCD), single-domain mild cognitive impairment (SD-MCI), multi-domain mild cognitive impairment (MD-MCI) and Alzheimer’s disease (AD). Volumes were standardized (z-scored) using the mean and standard deviation of the SCD group to facilitate comparison of atrophy rates across regions of different absolute sizes. Lines connect the mean volumes for each group across regions. A significant interaction between diagnostic group and brain region was observed, indicating that regional atrophy patterns differ by disease stage.

Significant group differences in thalamic volume were also observed across the groups (χ² = 44.2, df = 3, p < 0.001, **Figure 3C**). The AD group had smaller thalamus volumes compared to the MD-MCI (Z = -4.0, p_FDR_ < 0.001), SD-MCI (Z = -4.8, p_FDR_ < 0.001), and SCD (Z = -6.3, p_FDR_ < 0.001) groups. Furthermore, the MD-MCI group had smaller volumes than SCD group (Z = -3.9, p_FDR_ < 0.001). A positive correlation was also observed between total hypothalamic volume and thalamic volume (rho = 0.57, p_FDR_ < 0.001, **Figure 3D**), strongest in the anterior regions. For correlations between hypothalamic subregions volumes and total thalamic volume, see Supplementary Figure 9.

In comparison to the hypothalamus however, these group differences in hippocampal and thalamic volumes exhibited greater variability across the cognitive groups. Hypothalamic volume loss was less pronounced than both hippocampal and thalamic atrophy in both MD-MCI (interaction β = +0.025, p < 0.001) and AD (interaction β = +0.080, p < 0.001), suggesting that hippocampal and thalamic degeneration is disproportionately greater than hypothalamic decline in these groups (**Figure 3E**).

#### Association between hypothalamus volume and sleep quality

##### Subjective sleep

Across the whole sample, we found small positive correlations between PSQI global score and the whole hypothalamus (left: rho = 0.11, p_FDR_ = 0.010; right: rho = 0.18, p_FDR_ < 0.001). Sleep latency was associated with entire hypothalamus (left: rho = 0.10, p_FDR_ = 0.019; right: rho = 0.15, p_FDR_ < 0.001). For sleep duration, compared with individuals reported more than 7 hours sleep (β = 0.0539, 95% CI: (0.0532, 0.0546)), hypothalamus volume was larger in people with 5-6 hours sleep (β = 0.0559, 95% CI: (0.0551, 0.0566), p = 0.001) and 6-7 hours (β = 0.0558, 95% CI: (0.055, 0.0566), p = 0.001). For sleep efficiency, compared with individuals reported more than 85% sleep efficiency (β = 0.0545, 95% CI: (0.0539, 0.0551)), hypothalamus volume was larger in individuals with less than 65% sleep efficiency (β = 0.0565, 95% CI: (0.0554, 0.0577), p = 0.01) and those with 75%-84% sleep efficiency (β = 0.0559, 95% CI: (0.0549, 0.0569), p = 0.04). There were no associations between hypothalamus volume and scores of sleep quality (Component 1), medication (Component 6), or daytime function (Component 7). Exploratory associations between PSQI components and all hypothalamus subregions are detailed in the Supplementary materials.

We repeated the analyses excluding the AD group to mitigate potential confounding factors in reporting ability, yet positive correlations between PSQI global score and the whole right hypothalamus remained (rho = 0.15, p_FDR_ < 0.001, Supplementary Figure 10).

##### Objective sleep

The first 5 principal components (PC) explained the most variance in PSG measures (**Figure 4)**. The first principal component (PC1), related to obstructive sleep apnoea (OSA), explained 29% of the total variance; the second principal component (PC2), related to insomnia symptoms (sleep efficiency and sleep latency), explained 18.1% of the total variance; the third principal component (PC3), related to the amount of NREM sleep, especially deep SWS (NREM3), and explained 13.5% of the total variance; the forth principal component (PC4), also related to NREM sleep, but more towards light sleep (NREM1 and NREM2), explained 10.9% of the total variance; and the fifth principal component (PC5), related to amount of REM sleep and REM latency, and explained 9.8% of the total variance in the data (**Figure 4**).

**Figure 4.**
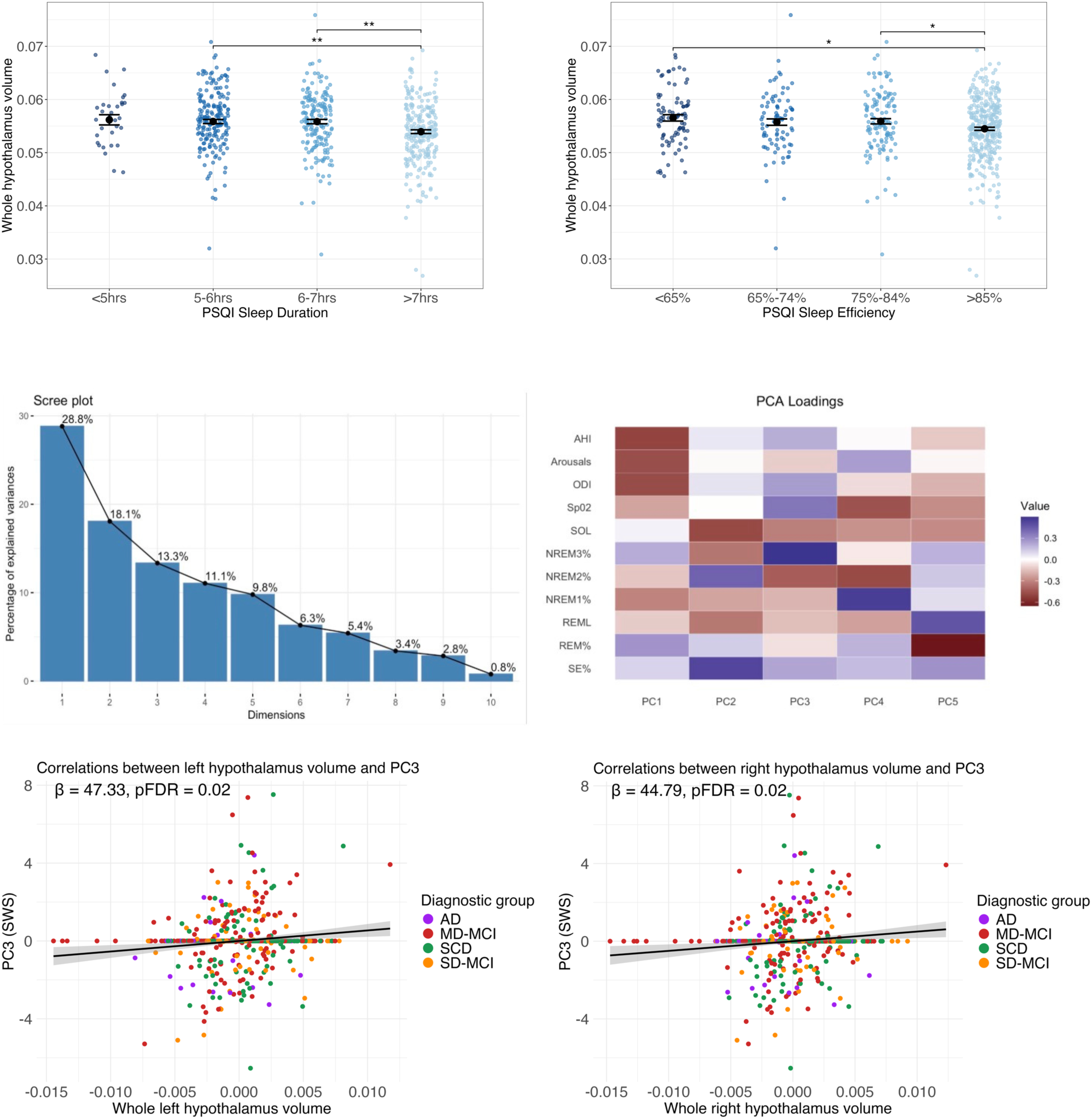
Hypothalamus volume is related to slow wave sleep. *Top Left:* Whole hypothalamic volume differences across different PSQI sleep duration across whole sample. *Top Right:* Whole hypothalamic volume differences across different PSQI sleep efficiency across whole sample. *Middle Left*: Scree plot of the explained variance explained by each principal component. The first 5 components were selected. *Middle Right:* Loadings of each PSG variable on each principal component (PC). *Bottom Left*: Correlation between left hypothalamic volume and PC3. *Bottom Right*: Correlation between left and right hypothalamic volume and PC3. All correlations were corrected for multiple comparisons using the FDR method.

To explore the relationship between sleep architecture and hypothalamic volume, we examined correlations between PCs 1-5 and hypothalamic volume. Across the whole sample, significant correlations were observed between PC3 (slow wave sleep) and whole left (β = 55.4, SE = 16.6, t = 3.3, pFDR = 0.009) and whole right (β = 50.8, SE = 16.6, t = 3.1, pFDR = 0.01) hypothalamus volume. There were no correlations between other PCs and volume total left or right hypothalamus in the whole sample.

In diagnostic groups, different associations were found between volume of hypothalamus subregions and PCs:

SCD group: PC4 (light sleep) was negative correlated with volume of anterior superior (β = - 385.1, SE = 149.3, t = -2.6, pFDR = 0.053). This correlation approached significance after FDR correction. This correlation was mainly drive by the right side (β = -735.2, SE = 263.3, t = -2.8, pFDR = 0.03).

SD-MCI group: A negative correlation was found between PC5 (REM sleep) and volume of anterior superior (β = -429.4, SE = 165.4, t = -2.6, pFDR = 0.03), anterior inferior (β = -479.1, SE = 181.3, t = -2.6, pFDR = 0.03), tubular superior (β = -145.1, SE = 63.3, t = -2.3, pFDR = 0.04).

MD-MCI group: PC3 (slow wave sleep) was positively associated with volume of anterior superior (β = 369.3, SE = 151.7, t = 2.4, pFDR = 0.02), anterior inferior (β = 400.9, SE = 153.1, t = 2.6, pFDR = 0.02), tubular superior (β = 112.7, SE = 43.4, t = 2.6, pFDR = 0.02), tubular inferior (β = 92.2, SE = 39.7, t = 2.3, pFDR = 0.03), and posterior (β = 98.2, SE = 39.0, t = 2.5, pFDR = 0.02) subregions.

AD group: There were no significant associations between objective sleep architecture and whole hypothalamus volume in the AD group. However, compared with the SCD group (β = 42.7, pFDR = 0.92), there were significantly stronger negative associations between PC2 (insomnia symptoms) and left anterior superior in the AD group (β = -2251.1, pFDR = 0.048). There is also a significant negative correlation between PC2 and volume of the tubular superior region in AD group (SCD: β = -19.9, pFDR = 0.96; AD: β = -424.8, pFDR = 0.048).

There were no other significant differences in correlations between the components and volume of either the whole left or right hypothalamus across diagnostic subgroups. Associations between hypothalamus volume and individual PSG metrics are reported in the Supplementary Materials (Supplementary Figures 11-13).

We also examined the correlations between volume of different hypothalamus subregions and PCs in different diagnostic groups based on amnestic symptoms. However, no correlations were observed between volume hypothalamus subregions and PCs in aMCI and naMCI group (all p>0.05). In addition, no correlations were observed between hippocampal or thalamic volume and objective sleep across the whole sample or within cognitive groups.

#### Hypothalamus volume and neuropsychological performance

We examined correlations between hypothalamus volume and neuropsychological performances:

*Verbal memory:* Verbal memory was positively associated with bilateral hypothalamus (left: rho = 0.14, p_FDR_ < 0.001; right: rho = 0.12, p_FDR_ = 0.003), anterior superior (rho = 0.19, p_FDR_ < 0.001), anterior inferior (rho = 0.13, p_FDR_ = 0.001), tubular superior (left: rho = 0.12, p_FDR_ = 0.002) and posterior (left: rho = 0.12; p_FDR_ = 0.003)

*Executive function:* Executive function was positively associated with bilateral hypothalamus (left: rho = 0.10, p_FDR_ = 0.013; right: rho = 0.08, p_FDR_ = 0.041), anterior superior (rho = 0.14, p_FDR_ < 0.001), anterior inferior (rho = 0.11, p_FDR_ = 0.007), and tubular superior (left: rho = 0.09, p_FDR_ = 0.033).

*Processing speed:* Processing speed was positively related to left hypothalamus (rho = 0.12, p_FDR_ = 0.002). As for subregion, it is weakly correlated with left anterior superior (rho = 0.1, p_FDR_ = 0.008), anterior inferior (rho = 0.1, p_FDR_ = 0.019), and posterior (rho = 0.09, p_FDR_ = 0.028).

#### Moderating role of hypothalamus volume in the relationship between hippocampus and verbal memory

We sought to explore if hypothalamus volume moderates the association between hippocampus volume and memory performance. Across the whole sample, the hypothalamic volume moderated the effect of hippocampal volume on verbal memory (β = -0.07, SE = 0.03, t = -2.1, p = 0.04) (see Figure 5). This effect was mainly driven by the anterior superior region (β = -0.10, SE = 0.03, t = -2.9, p = 0.004).

**Figure 5.**
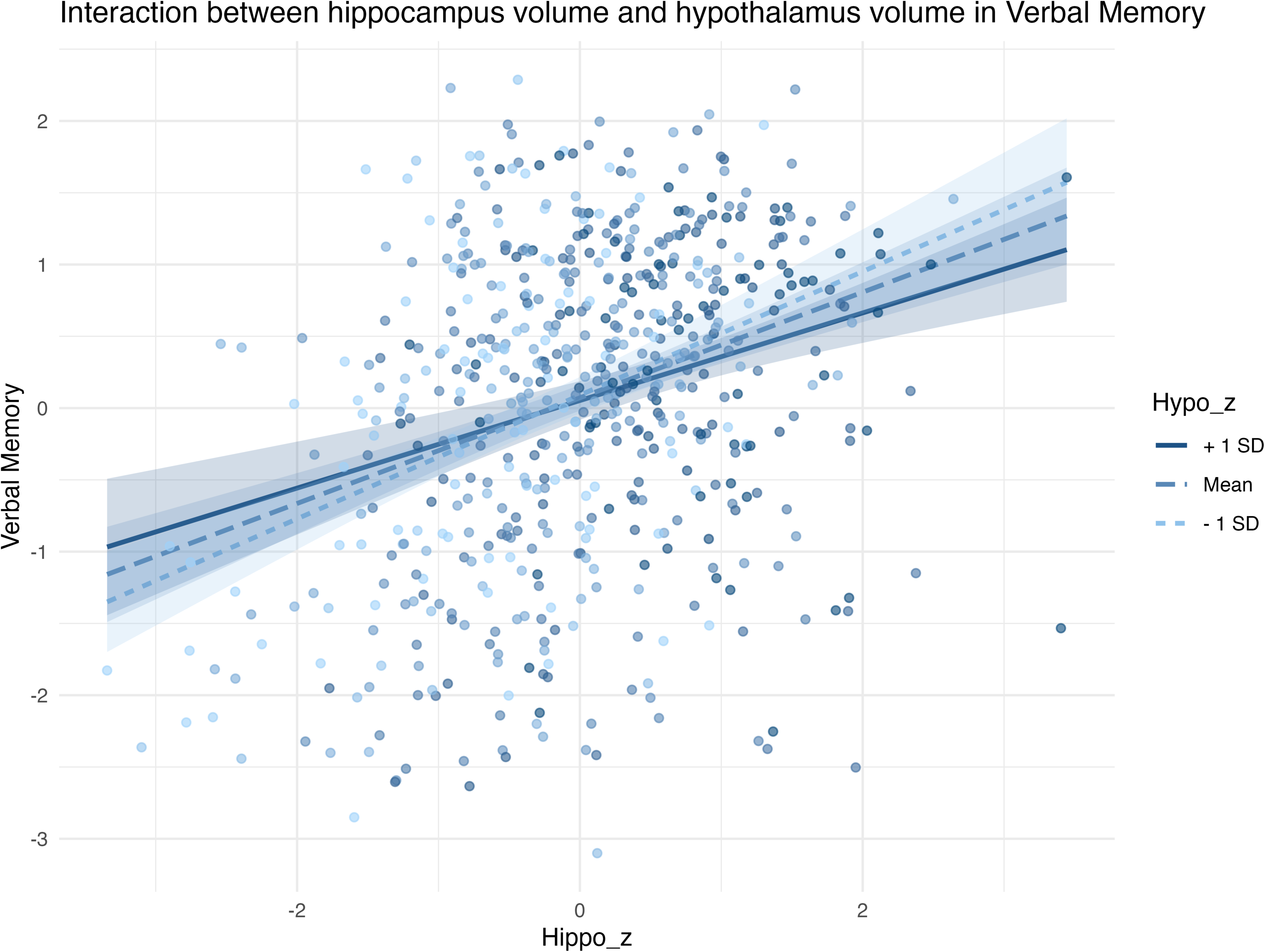
Hypothalamus volume moderates the relationship between hippocampus volume and verbal memory. Moderation scatter plot between hippocampus volume: hypothalamus volume and verbal memory. All correlations were corrected for multiple comparisons using the FDR method. Hypo = hypothalamus; Hippo = hippocampus.

## Discussion

This study provides evidence that the hypothalamus differs systematically across the Alzheimer’s disease continuum and represents a relevant node linking sleep disruption and cognitive impairment. We observed clear and regionally specific differences in hypothalamic volume across the Alzheimer’s disease (AD) clinical continuum, with the most pronounced reductions localised to anterior subregions and evident already in individuals with probable AD and multiple-domain MCI but not yet observed in the single-domain MCI stage. These findings align with previous reports of hypothalamic involvement in AD and demonstrate that anterior hypothalamic differences are detectable across intermediate stages of cognitive impairment, not only in established dementia.[20,21] The pattern is consistent with earlier work highlighting disproportionate anterior involvement and suggests that these subregions may represent a selectively vulnerable node within the broader AD-affected network and warrants further investigation as a potential site of Alzheimer’s disease progression.[22]

While hypothalamic volume was smaller in MD-MCI and AD groups, the extent of this reduction was less marked than that observed in the hippocampus or thalamus. These findings suggest that hypothalamic involvement in neurodegeneration may lag behind or be less pronounced than hippocampal and thalamic atrophy in the progression of Alzheimer’s disease. Nevertheless, we observed that hypothalamic integrity may play a specific role in sleep disturbances observed in MCI and AD. Consistent with our hypothesis, we found that lower volume of the bilateral hypothalamus was associated with less slow wave sleep across the whole sample. Positive correlations were also observed with REM and slow wave sleep in MCI groups. These correlations were primarily observed in the anterior subregions and tubular superior regions, followed by posterior regions and tubular inferior. Meanwhile, the relationships between hypothalamic subregion volumes and other sleep parameters varied across different groups. Specifically, compared to the SCD groups, the AD groups exhibited a relatively stronger negative correlation between hypothalamic subregion volumes and insomnia symptoms (lower sleep efficiency and longer sleep latency). These differences were primarily observed in the superior regions of the hypothalamus.

Our observations suggest that while the hippocampus and thalamus were relatively more impacted by the progression of AD and associated cognitive decline, the hypothalamus was more sensitively associated with objective sleep parameters, particularly reduced slow wave sleep. This reinforces the specific dominance of the hypothalamus in sleep disturbance. The lack of relationships between sleep and the hippocampus may reflect asynchronous pathological progression between the hippocampus and hypothalamus or indicate that Aβ accumulation in the hypothalamus may be more directly influenced by sleep disturbances than the degeneration of hippocampus. These findings support the hypothesis that atrophy of the hypothalamus may contribute to significant changes in sleep quality and architecture in the continuum of cognitive decline and neurodegeneration.

There have been very few studies exploring the relationship between hypothalamic structure and objective sleep, particularly in the context of Alzheimer’s disease. A study in Huntington’s disease reported no association between hypothalamic volume and sleep measures [53], whereas more recent work has identified positive associations between REM sleep proportion and anterior hypothalamic volume in middle-aged people with OSA [54], consistent with our findings in MD-MCI group. Experimental post-mortem studies further demonstrate that neuronal loss within sleep-regulatory hypothalamic nuclei, including the ventrolateral preoptic nucleus (VLPO) and suprachiasmatic nucleus (SCN), are associated with sleep disruption [17,55,56], with VLPO lesions leading to reduction in REM and NREM sleep time in animal models [17]. Taken together, these findings provide a biological context for our observation that structural differences in anterior hypothalamic subregions are associated with alterations in REM and NREM sleep and support the view that hypothalamic integrity contributes to sleep architecture changes in early neurodegenerative disease.

Previous studies have suggested an association between slow-wave sleep (SWS) and amyloid-β burden [57]. Amyloid-β accumulates during wakefulness, in part due to increased oxidative stress and reduced perivascular clearance [58,59], whereas SWS has been proposed to facilitate clearance through opposing mechanisms, including enhanced glymphatic activity [59,60]. In this context, the observed association between reduced hypothalamic volume and diminished SWS is consistent with a potential role for hypothalamic integrity in sleep-related clearance processes. However, this interpretation remains indirect, and future studies directly assessing hypothalamic involvement in perivascular or glymphatic function are required to substantiate this link.

Interestingly, compared with the SCD group, more sleep latency and less sleep efficiency were correlated with larger left anterior superior and tubular superior volume in the AD group. Previous studies have implicated hypothalamic dysfunction in AD patients with the presence of sleep disorders [11,61]. This counterintuitive finding cannot be explained with volumetric changes alone. We hypothesise that it may either reflect an inflammation response in AD, or widespread disruption of neuroendocrine signalling during advanced disease stage, which results in the disabling of the remaining neurons [62].

Regarding the correlations between hypothalamus volume and subjective sleep quality, surprisingly, we found a weak positive correlation between PSQI scores and hypothalamus volume, which suggests larger hypothalamus volume is related to worse subjective sleep quality. To date, there has been little research revealing the association between subjective sleep quality and hypothalamus volume. A recent study reported a positive correlation between hypothalamus volume and subjective sleep quality in healthy older people [63]. Ding et al. [64] suggested the resting-state functional connectivity (RSFC) between hypothalamus and medial prefrontal cortex (mPFC) is positively correlated with PSQI score. However, as we found that the AD group had lower scores on the PSQI (i.e. reported better subjective sleep quality), this highlights the difficulty of assessing the subjective experience of sleep in later stages of neurodegeneration, where cognitive decline (and thus recall) is most impacted [65,66].

Finally, hypothalamus volume was positively correlated with memory performance, executive functions and processing speed. In subregions, the volume of anterior region was positively related to all neuropsychological performance. The volume of tubular superior was positively associated with memory and executive function. Posterior volume was positively related to memory and processing speed. Overall, these findings likely reflect the fact that hypothalamus volume was most reduced in multiple-domain MCI and AD, the groups suffering the worst cognitive decline. Importantly however, the hypothalamus volume moderated the correlation between hippocampus volume and verbal memory, i.e., larger hippocampal volume was consistently associated with better memory performance, but this relationship was stronger in individuals with larger hypothalamic volumes. The moderation effect was mainly driven again by the anterior superior region, consistent with existing knowledge of the crucial role of PVN in memory [67,68]. As there are direct fibre projections from magnocellular neurons in the PVN to the hippocampus [69], along with reciprocal pathways projecting from the hippocampus back to the PVN [70], it is possible that memory deficits are driven by the interaction between these two regions. Consistent with recent findings [28], executive functions were mainly positively correlated with anterior subregions, apart from PVN, as these regions contain SCN and VLPO, which are explained above, involved in sleep regulation. Given the established link between sleep quality and executive function [71], this anatomical association may reflect the functional interplay between sleep-related neural circuits and cognitive performance.

This study has a number of strengths, including the measure of sleep parameters employing gold-standard objective PSG. This study also represents the first investigation into hypothalamic volume variations across the continuum of AD pathology (spanning from SCD to SD-MCI, MD-MCI and AD). Nonetheless, this study also has several limitations that should be acknowledged. Firstly, the absence of a healthy control group limits our ability to establish baseline comparisons and to assess deviations in sleep-related measures across different neurodegenerative stages. Secondly, the cross-sectional and correlational nature of the study precludes any causal interpretations regarding the directionality of the relationship between hypothalamic structure and sleep parameters, which should be followed up in a longitudinal sample. Although our volumetric approach offers a high degree of anatomical precision, the segmentation scheme used delineates the hypothalamus into five subregions rather than individual hypothalamic nuclei, which may obscure more localized effects. Additionally, our analysis was restricted to structural measurements of hypothalamic volume and clinical classifications of the AD continuum, therefore future studies would benefit from integrating multimodal biomarkers, including relevant neuroendocrine markers, signalling molecules, and Aβ burden, and connectivity pathways between the hypothalamus and hippocampus, to more comprehensively investigate the bidirectional influences between hypothalamic integrity, sleep regulation, and the pathophysiological progression of AD.

## Conclusion

This study revealed the atrophy pattern of the hypothalamus at different stages of AD progression, with the most pronounced atrophy occurring in the anterior subregions. These findings suggest that hypothalamic volume is associated with objective sleep quality, suggesting a significant role in mediating sleep disturbances in neurodegeneration - which can further impact cognitive decline. Future research should incorporate additional biomarkers to understand the bidirectional relationship between sleep and hypothalamic structure throughout the course of AD. Moreover, future interventional studies focused on orexinergic pathways may be important in improving sleep quality for reducing the risk of dementia.

## Data Availability

All data produced in the present study are available upon reasonable request to the authors

## Acknowledgments

We thank all the participants from the Healthy Brain Ageing Clinic who contributed their time to being a part of this study. We acknowledge the contribution of all the clinical and administrative staff members at the Healthy Brain Ageing Clinic, the Woolcock Institute of Medical Research, and the Brain and Mind Centre.

## Consent statement

All participants provided written informed consent.

