## Supplementary for "Hypothalamic structural differences link sleep and cognition across the Alzheimer’s disease spectrum"

Supplementary Materials

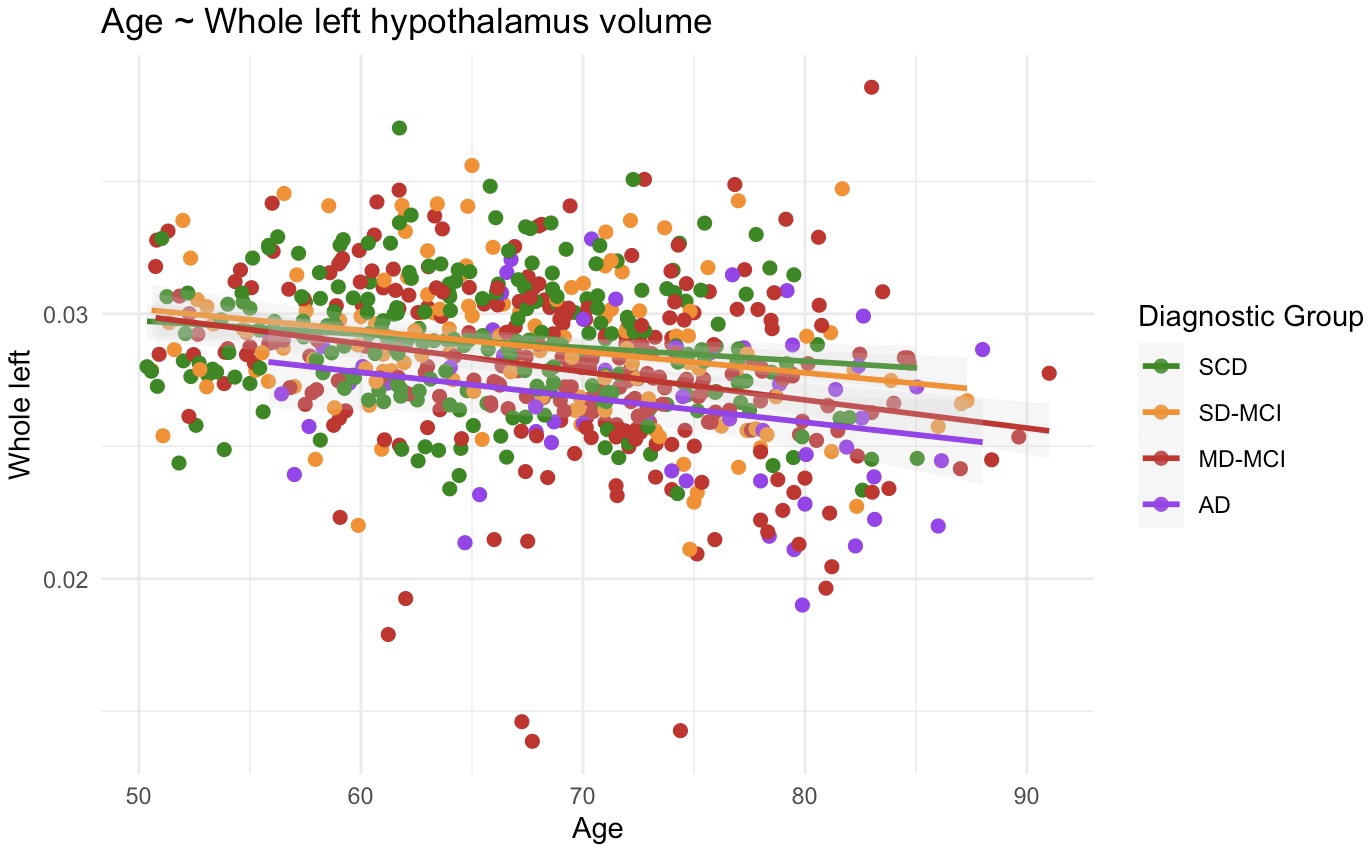

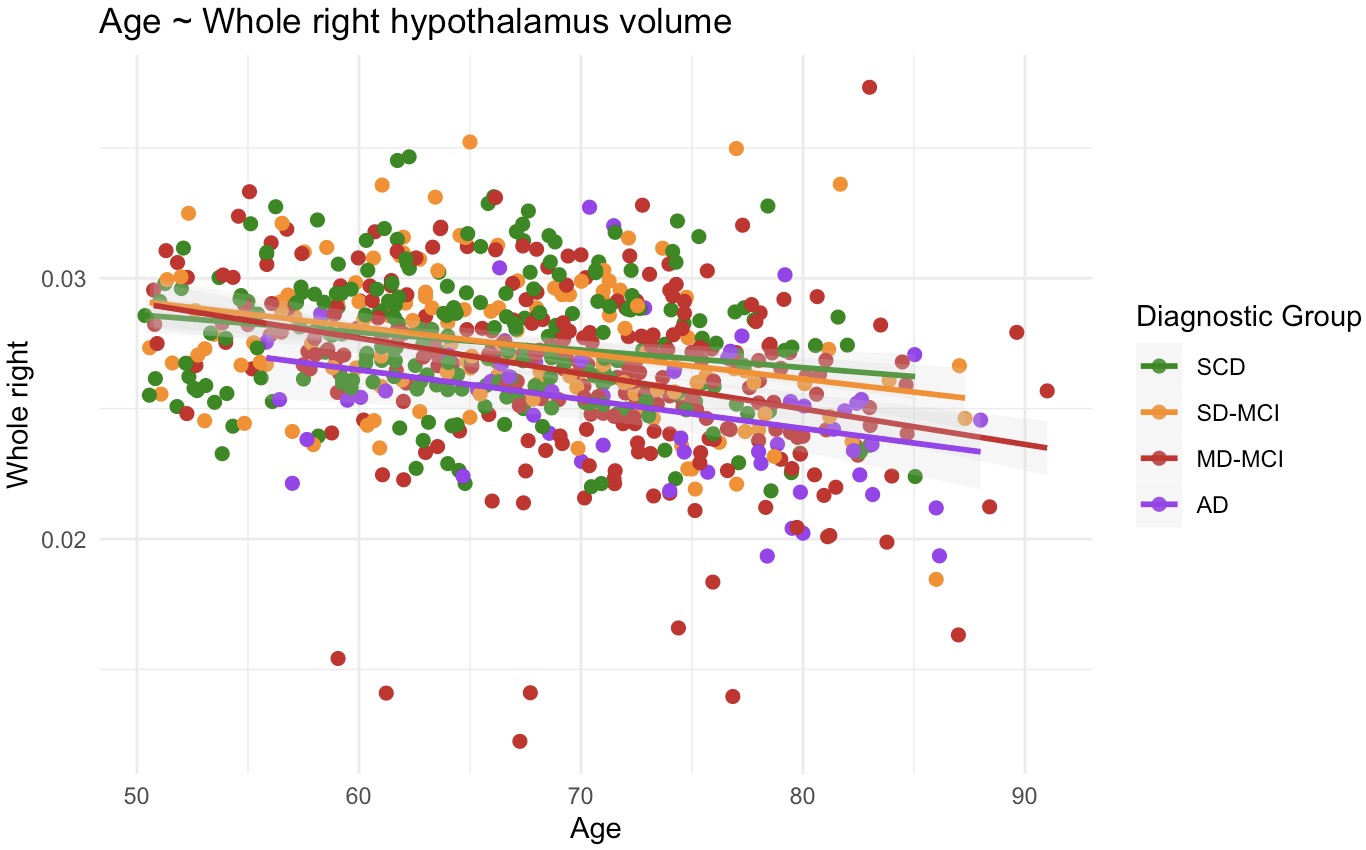

Supplementary Figure 1. The associations between age and left & right hypothalamus volume

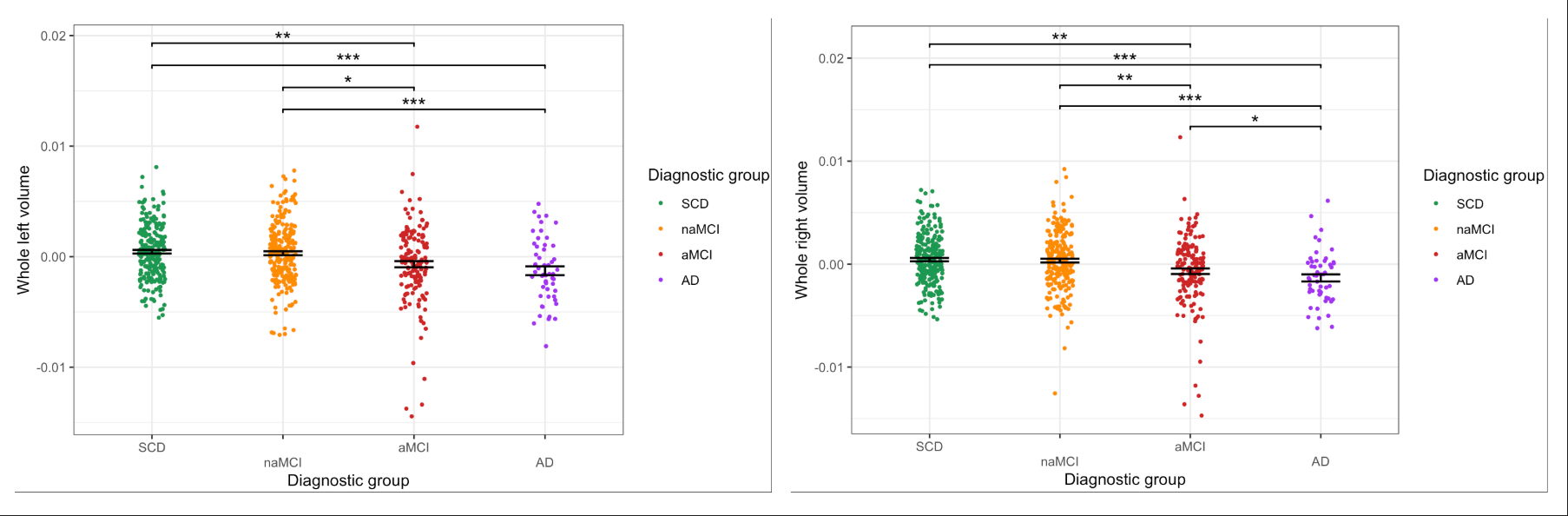

Supplementary Figure 2. The differences on whole hypothalamus volume across SCD, naMCI, aMCI and AD groups

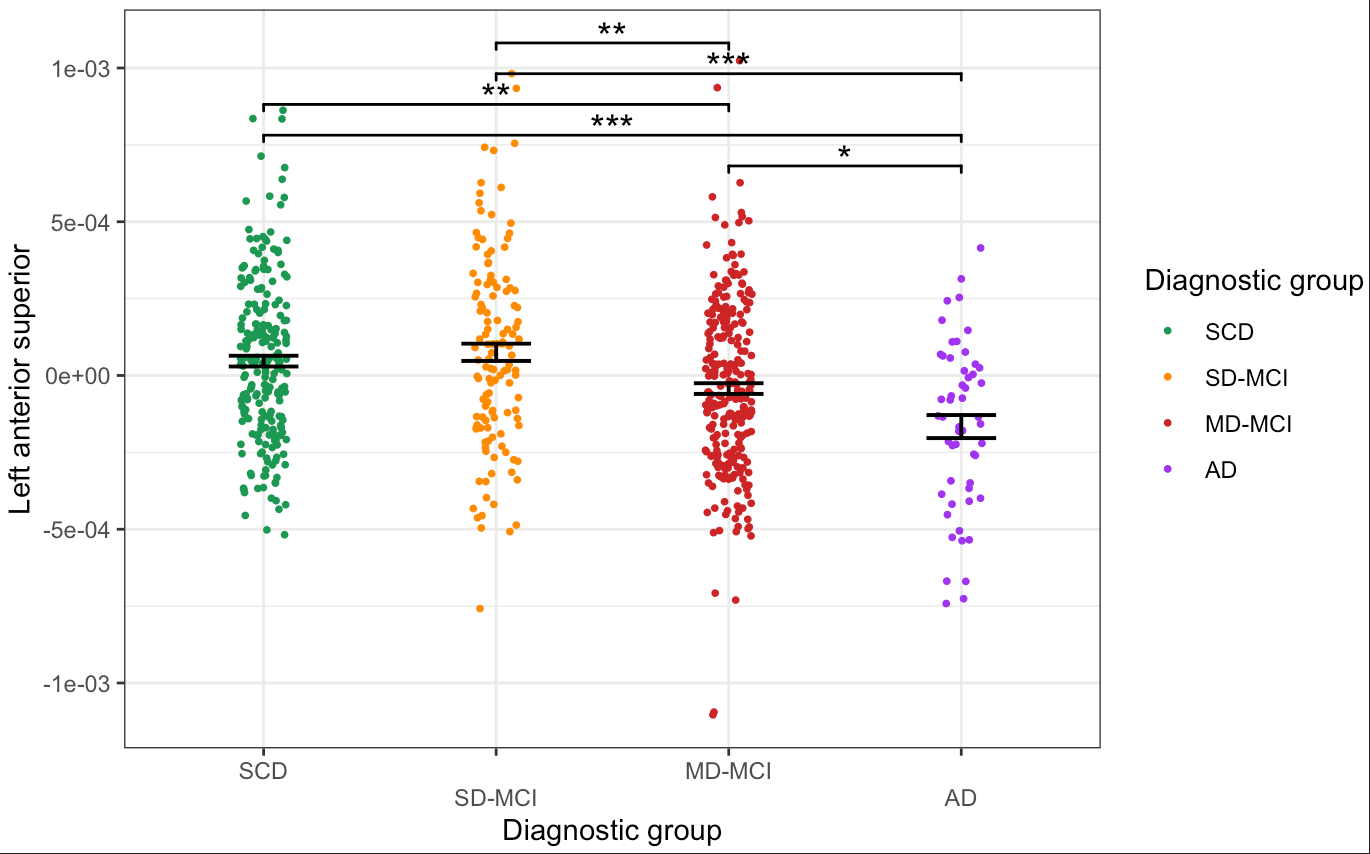

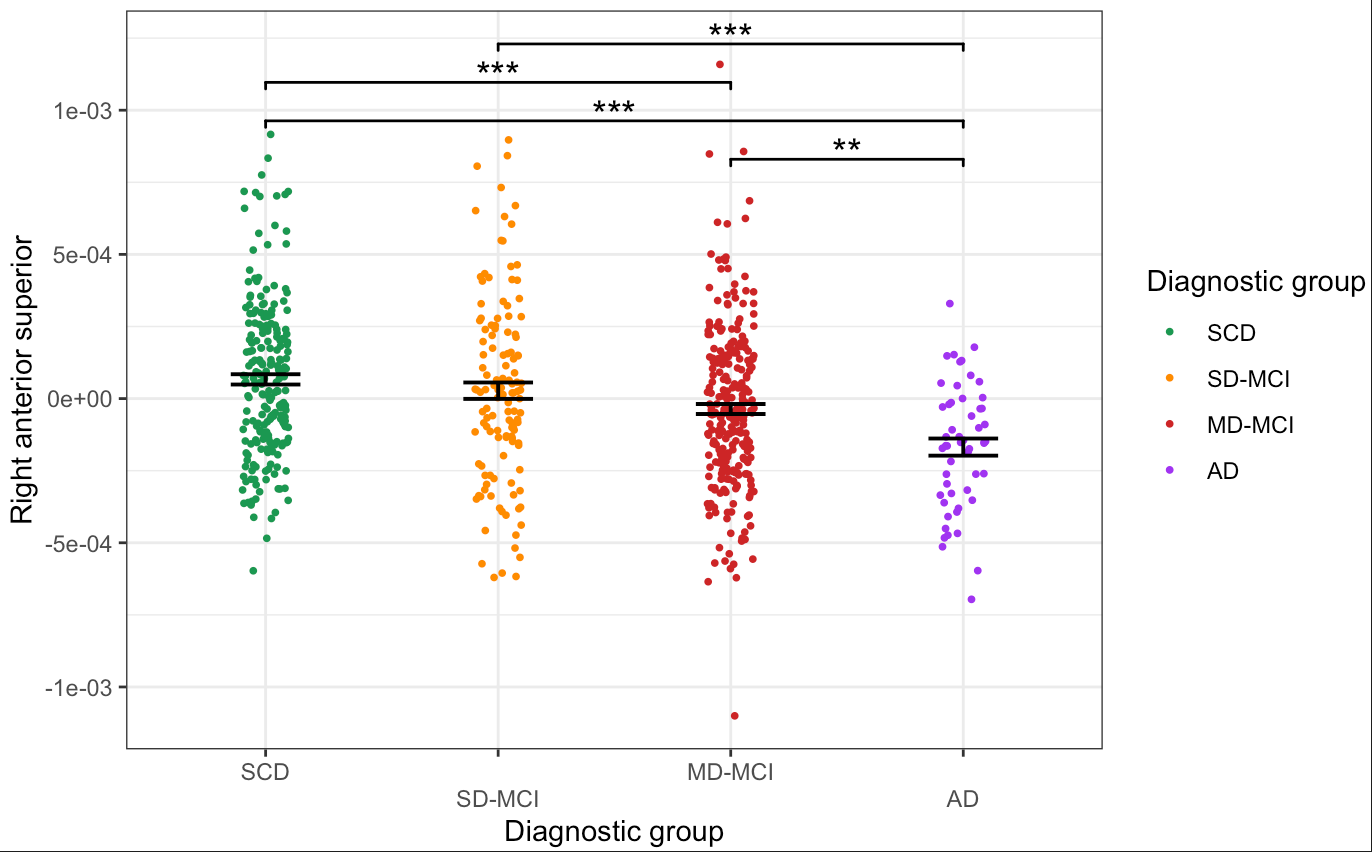

Supplementary Figure 3. The differences on left & right anterior superior hypothalamus volume across different diagnostic groups

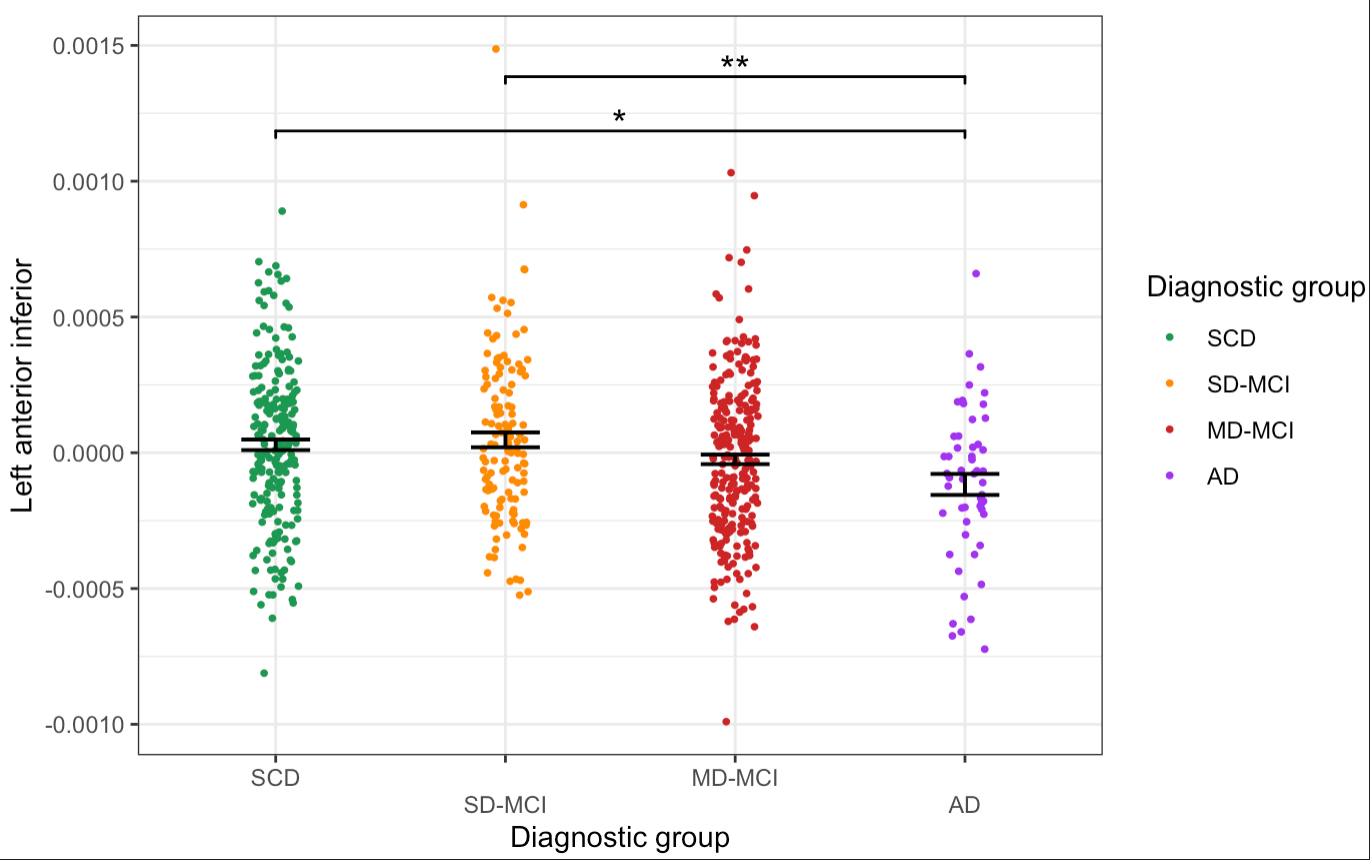

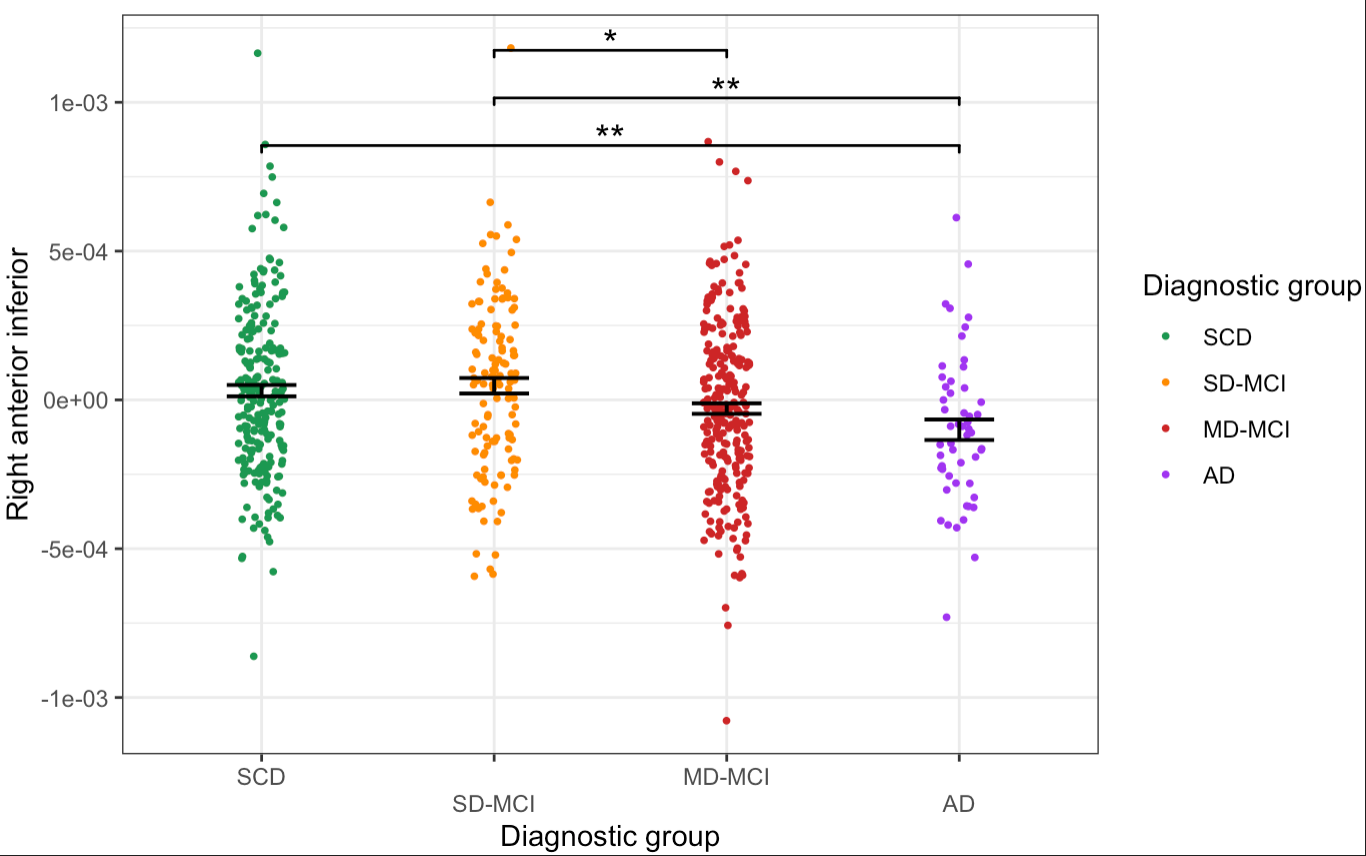

Supplementary Figure 4. The differences on left & right anterior inferior hypothalamus volume across different diagnostic groups

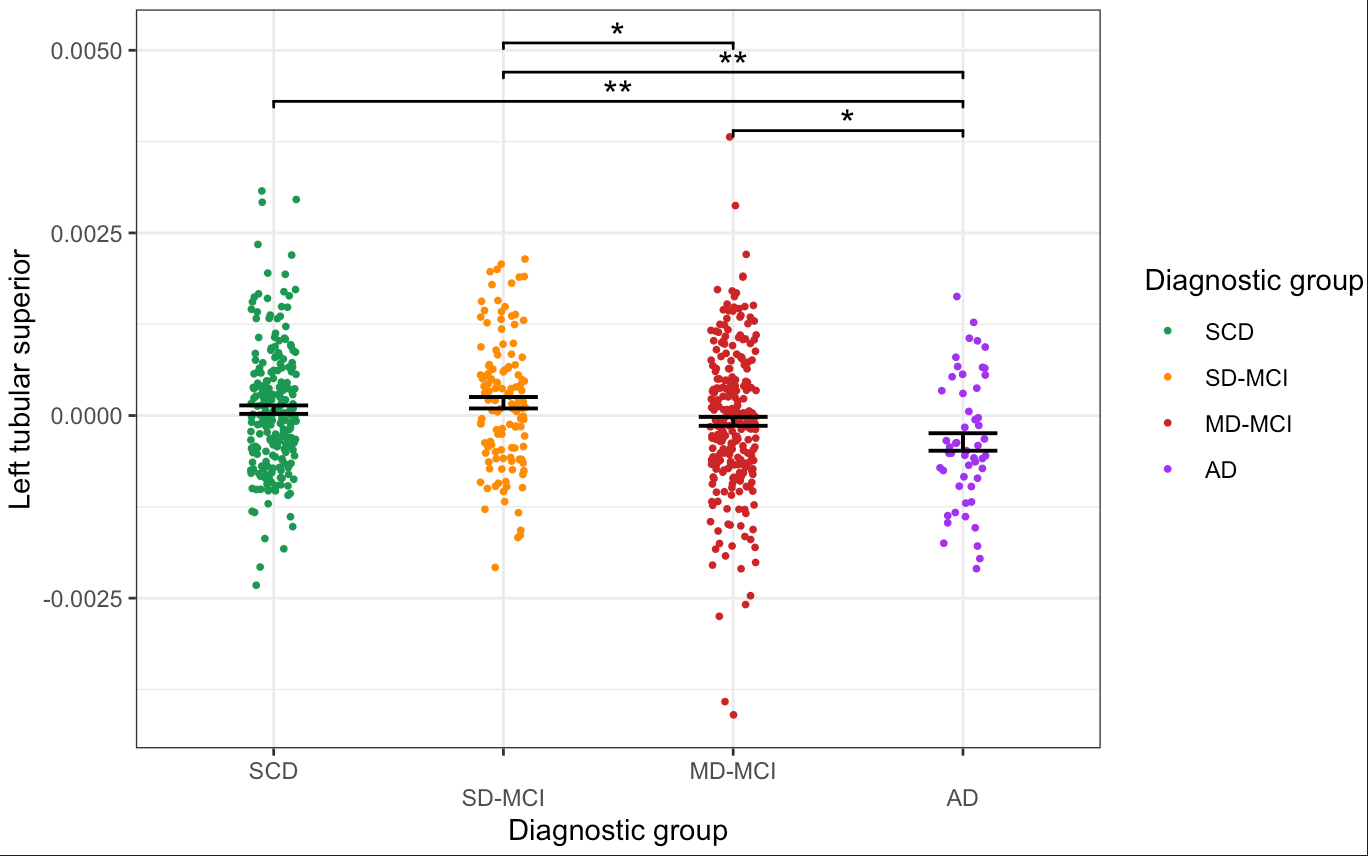

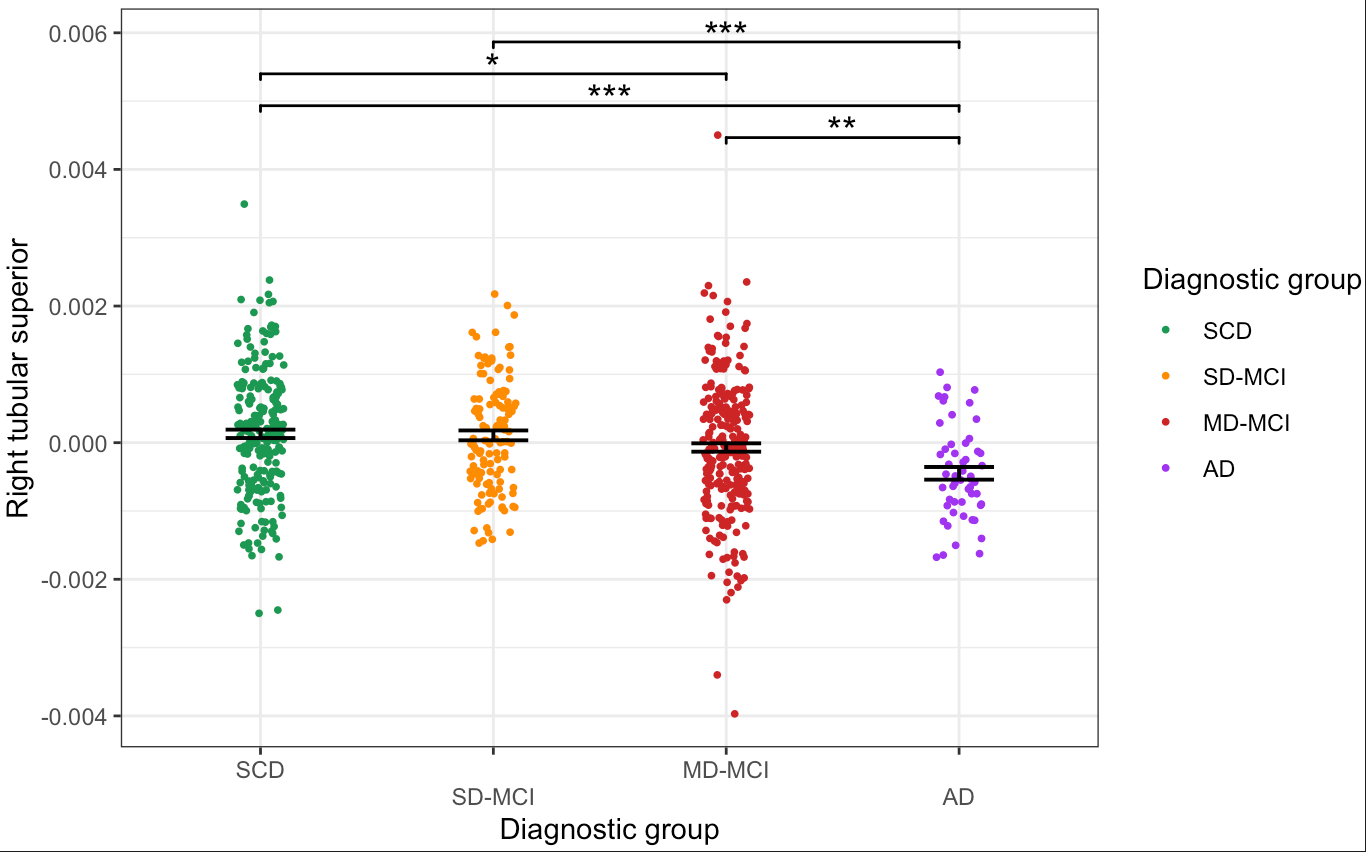

Supplementary Figure 5. The differences on left & right tubular inferior hypothalamus volume across different diagnostic groups

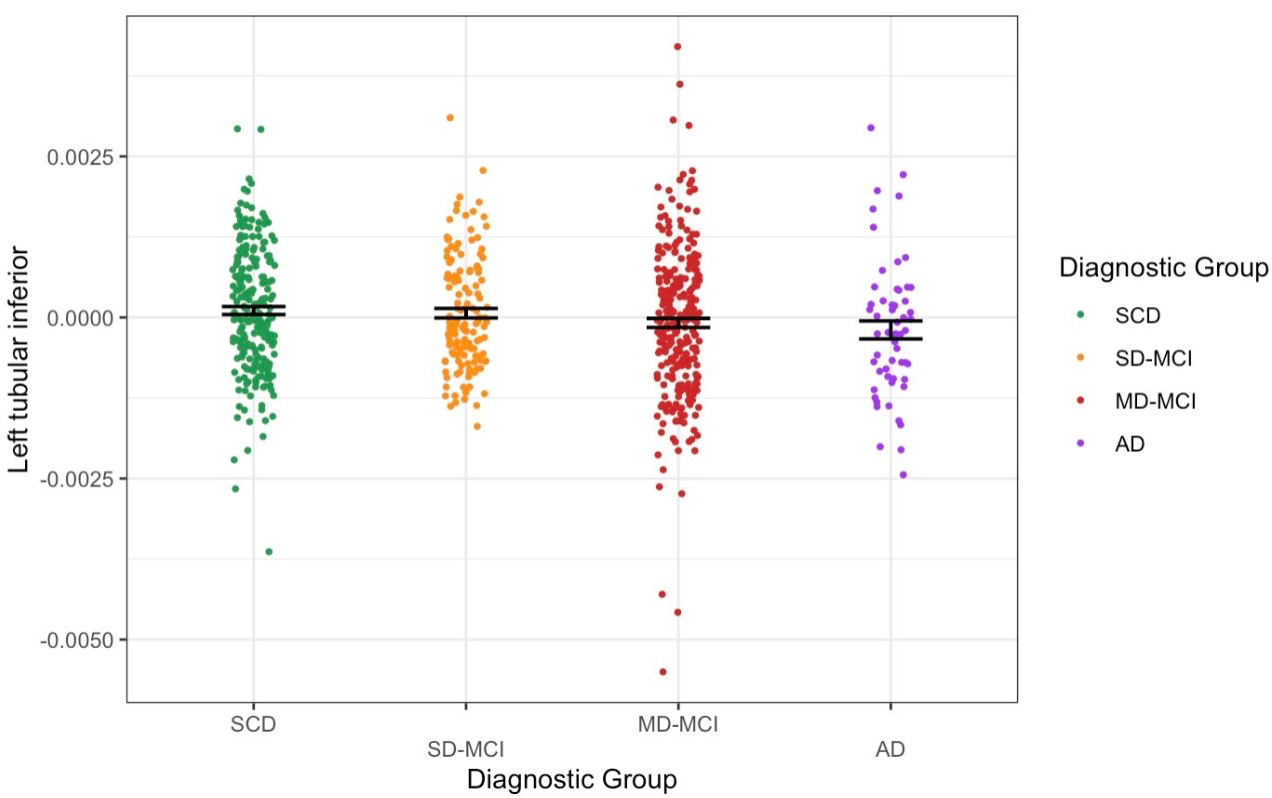

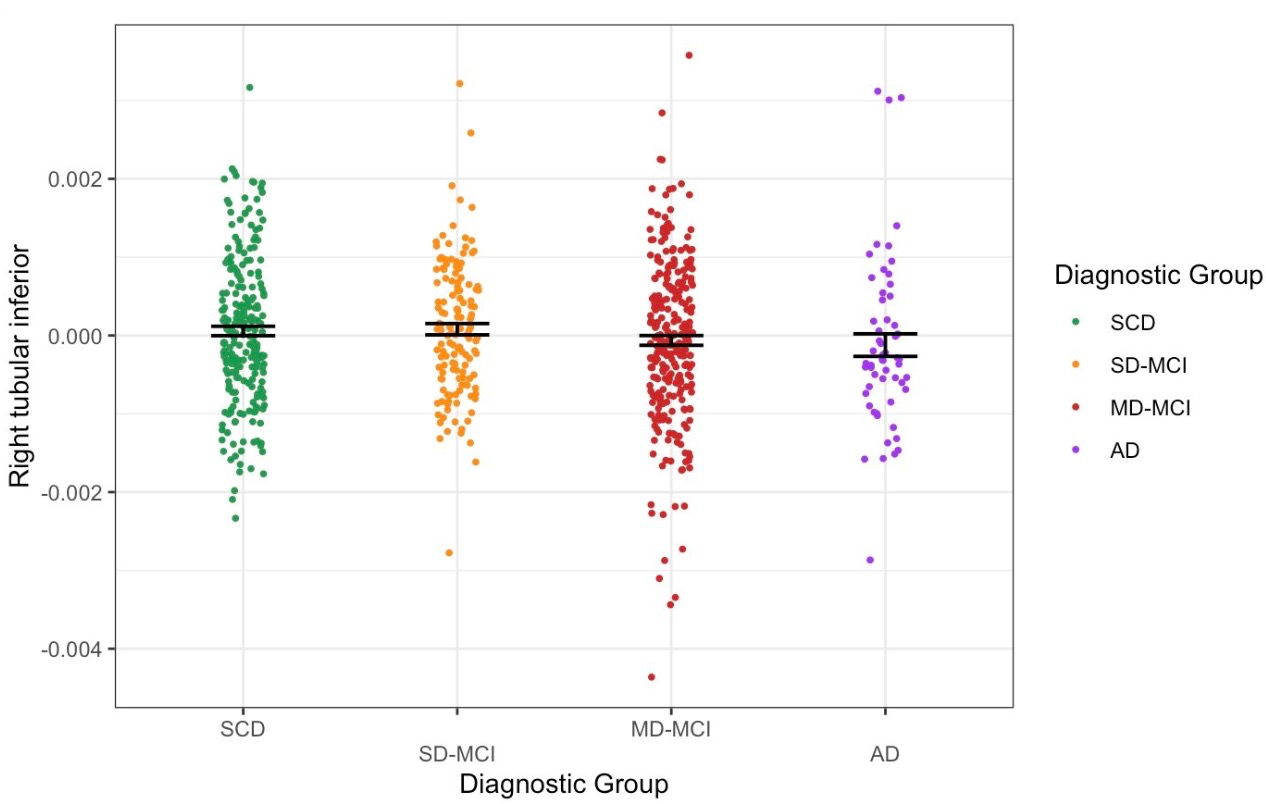

Supplementary Figure 6. The differences on left & right tubular superior hypothalamus volume across different diagnostic groups

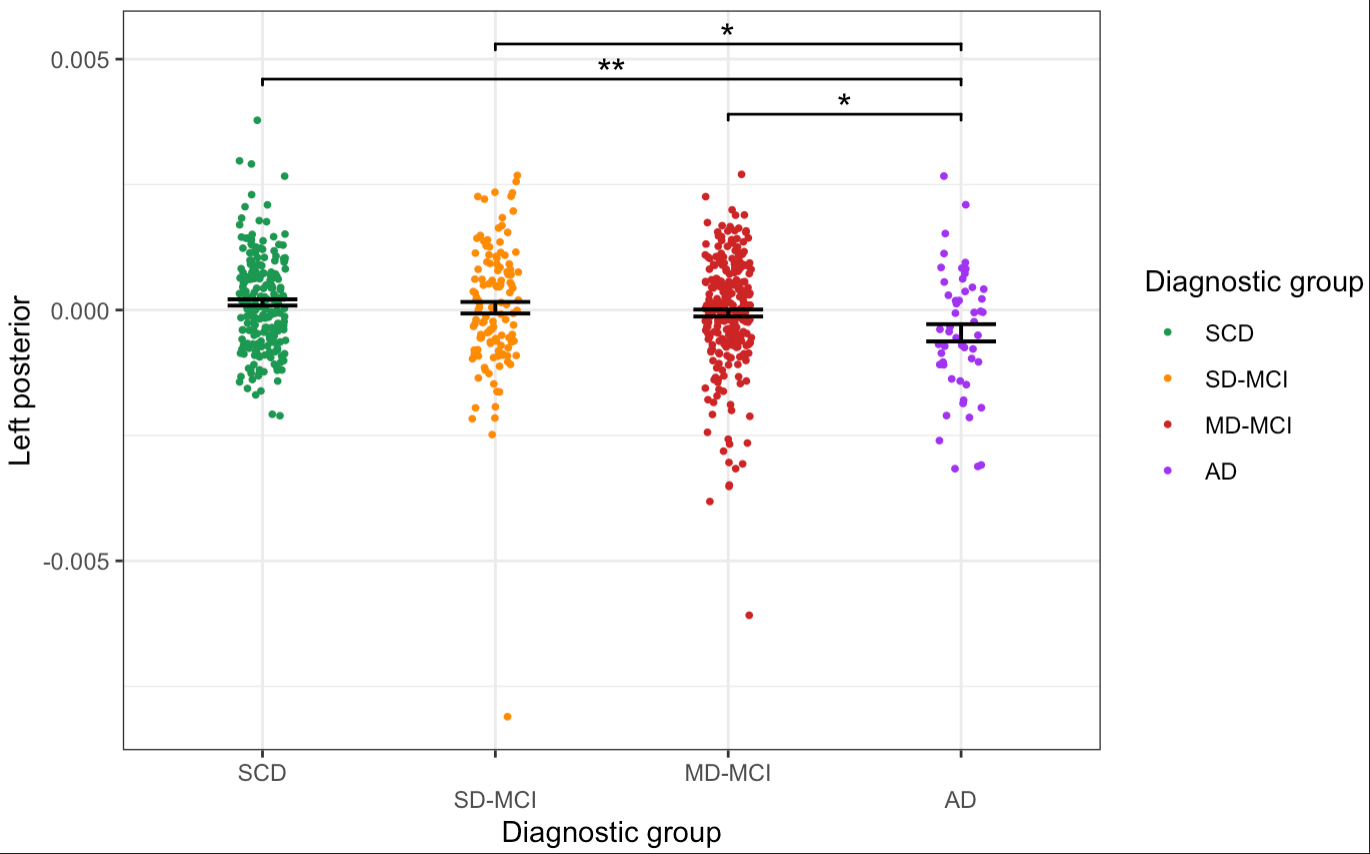

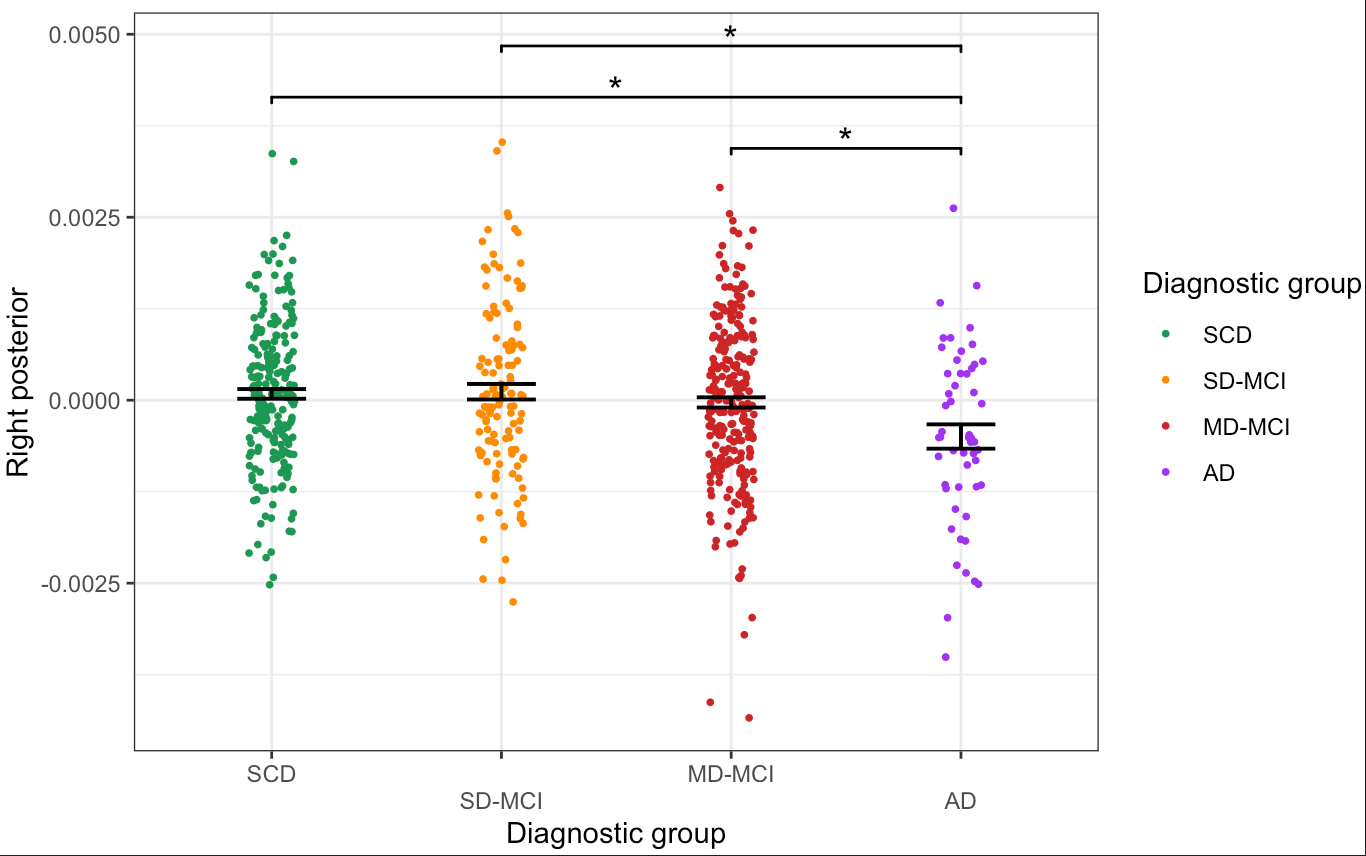

Supplementary Figure 7. The differences on left & right posterior hypothalamus volume across different diagnostic groups

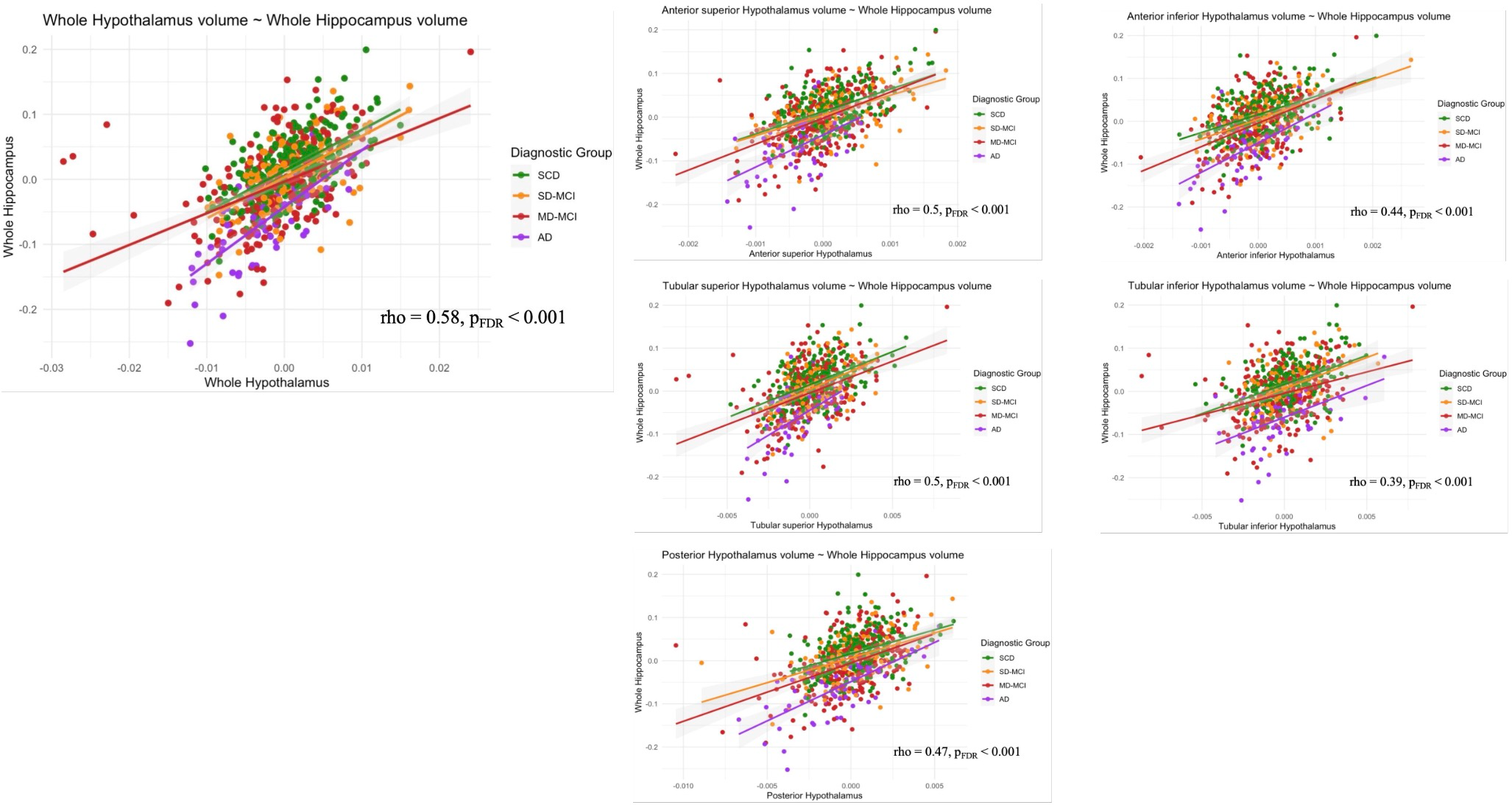

Supplementary Figure 8. The associations between hypothalamus volume and hippocampus volume

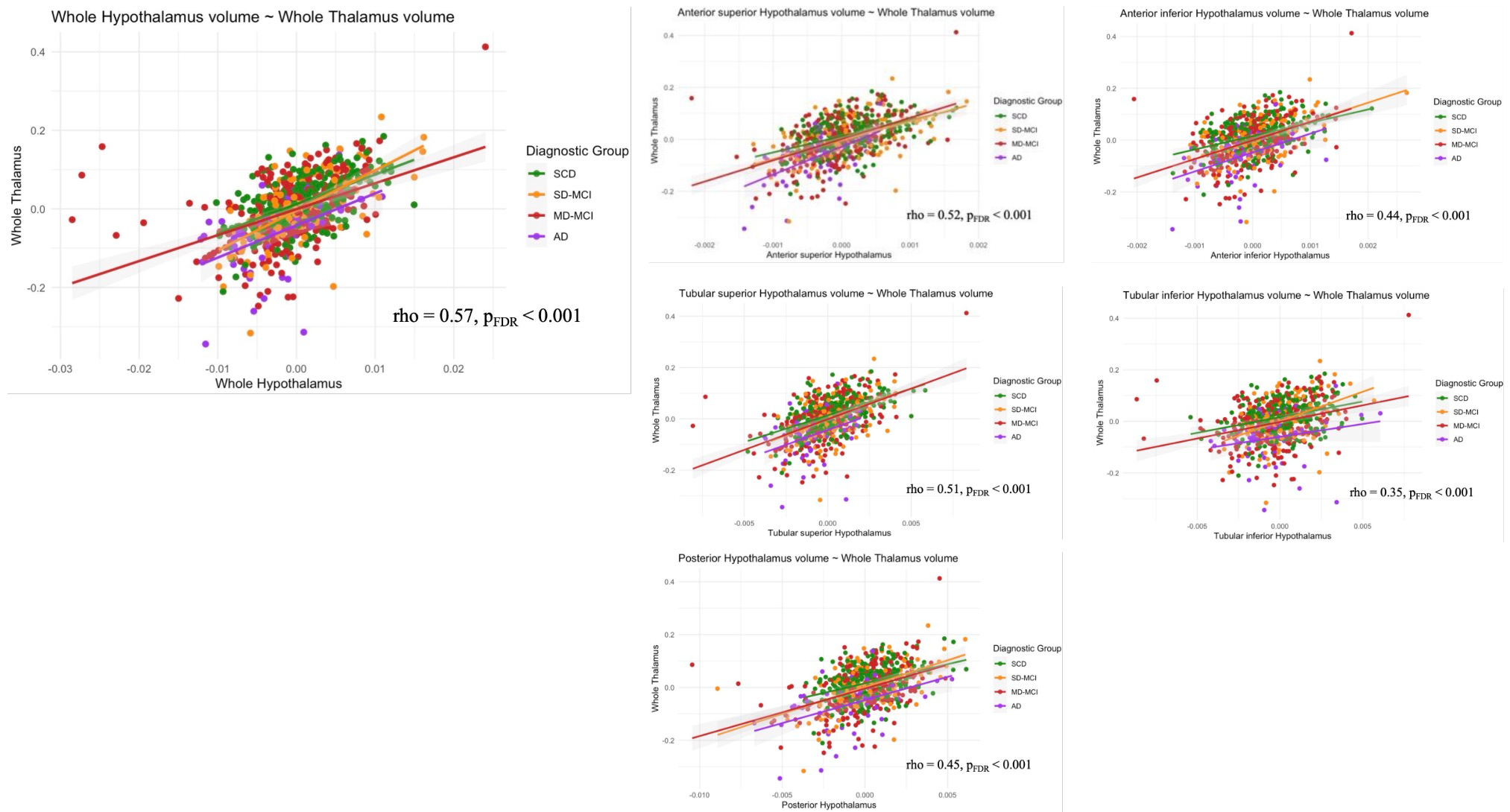

Supplementary Figure 9. The associations between hypothalamus volume and thalamus volume

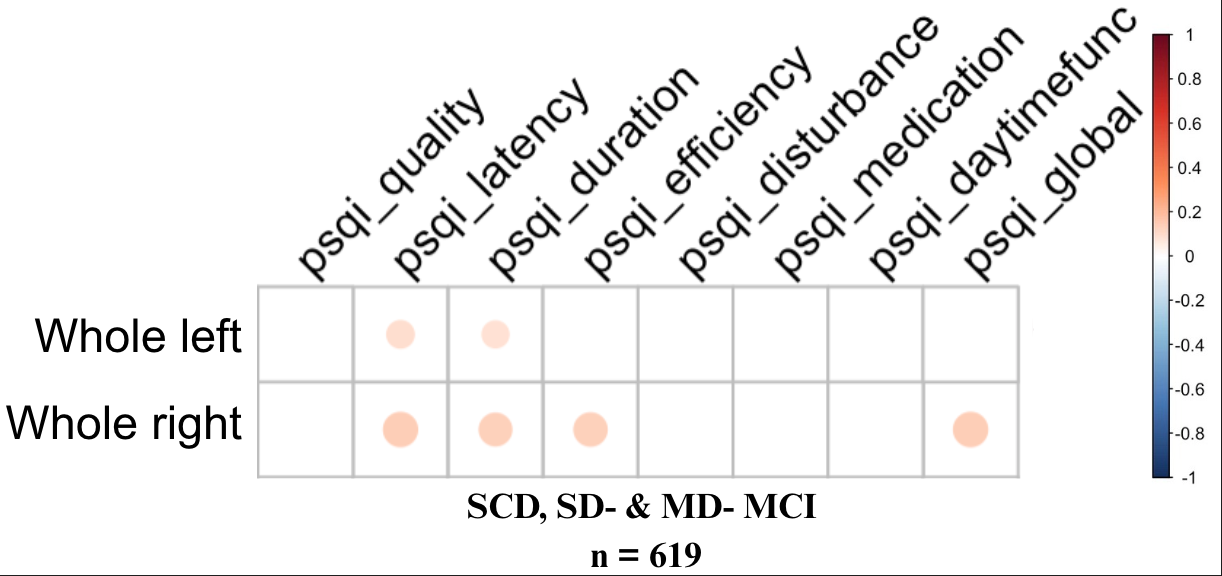

Supplementary Figure 10. The correlations matrix between PSQI and whole left&right hypothalamus volume

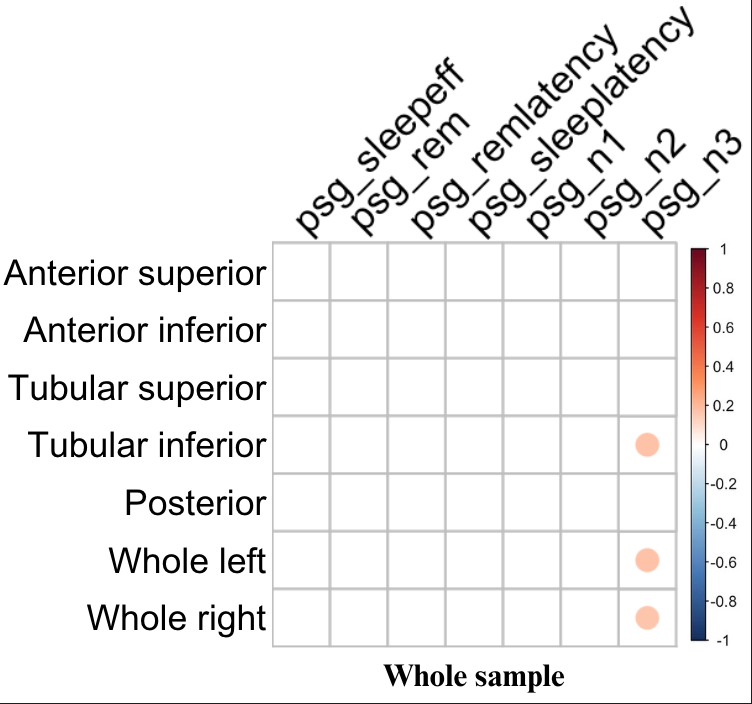

Supplementary Figure 11. The correlations matrix between PSG sleep parameters and hypothalamus volume across whole sample

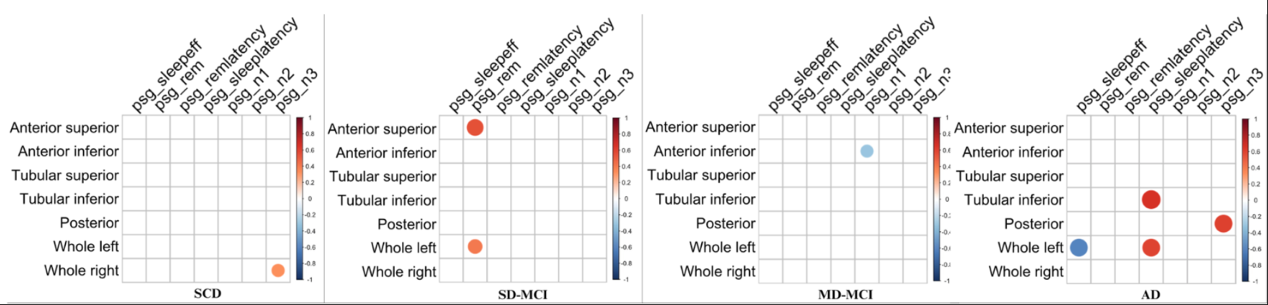
Supplementary Figure 12. The correlations matrix between PSG sleep parameters and hypothalamus volume in different diagnostic groups.

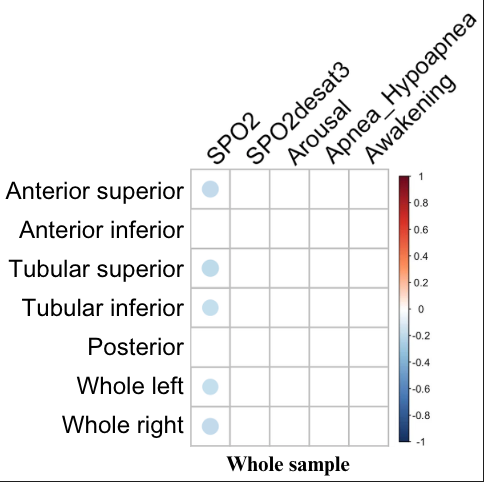

Supplementary Figure 13. The correlations matrix between OSA variables and hypothalamus volume across whole sample

**Supplementary Methods**

MRI acquisition parameters

For site 1, prior to July 2019, the structural MRI data were collected using a 3-Tesla GE Discovery MR750 scanner (GE Medical Systems, Milwaukee, WI) with an 8-channel phased-array head coil. For those scans acquired after July 2019, the structural MRI data were performed on a 32-channel phased-array head coil. For site 2, the structural MRI data were collected using a 3-Tesla Siemens Magnetom Skyra scanner (SIEMENS, Erlangen, Germany) with a 20-channel phased-array head coil. For those scans acquired with a 8-channel head coil, the following T1 weighted imaging sequence was used: 3D-T1-weighted BRAVO Spoiled Gradient-Recalled (SPGR) sequence (phase acceleration factor = 2), acquiring 196 sagittal slices (repetition time = 7.2 ms; echo time = 2.8 ms; flip angle = 12; matrix 256×256; 0.9 mm isotropic voxels), acquisition time = 4 minutes, 27 seconds. For scans obtained using a 20-channel head coil, The following T1 weighted imaging sequence was used: T1 MPRAGE sequence (acquiring 192 sagittal slices (repetition time = 2200.0 ms; echo time = 2.48 ms; flip angle = 8; matrix 248×256). For those one with a 32-channel head coil, the following 3D-T1-weighted BRAVO Spoiled Gradient-Recalled (SPGR) sequence was implemented: 176 sagittal slices (repetition time = 7.4 ms; echo time = 3.0 ms; flip angle = 11; matrix 256×256; 1.0 mm isotropic voxels), acquisition time = 4 minutes, 05 seconds.

**Supplementary Results**

Correction of channel of head coil

Specifically, on the left side, there was significant difference between SD-MCI and MD-MCI group (β = 0.00075, SE = 0.0003, t(666) = 2.5, p_FDR_ = 0.02). The difference between MD-MCI and AD was no longer significant but bordered on significance (β = 0.00083, SE = 0.00042, t(666) = 2.0, p_FDR_ = 0.057). On the right side, significant volume differences were found between SCD and MD-MCI group (β = 0.00068, SE = 0.00026, t(666) = 2.6, p_FDR_ = 0.02), and SD-MCI and MD-MCI group (β = 0.00071, SE = 0.0003, t(666) = 2.4, p_FDR_ = 0.03).

Volume differences across groups classified by onset of memory deficits
Based on the onset of memory impairment, MCI groups were classified as Amnestic MCI (aMCI) and Non-amnestic MCI (naMCI) groups, AD and SCD kept the same.

Both the left and right hypothalamus volume differed across SCD, SD-MCI, MD-MCI and AD groups (left: χ² = 23.4, df = 3, p < 0.001；right: χ² = 30.0, df = 3, p < 0.001)
In the left hypothalamus, significant differences were observed between AD and SCD (z = -4.0, p_FDR_ < 0.001), AD and naMCI (z = -3.6, p_FDR_ < 0.001), aMCI and SCD (z = -3.2, p_FDR_ = 0.003), aMCI and naMCI (z = -2.7, p_FDR_ = 0.01). Similar differences were observed in the right hypothalamus between AD and SCD (z = -4.5, p_FDR_ < 0.001), AD and naMCI (z = -4.5, p_FDR_ < 0.001), AD and aMCI group (z = -2.2, p_FDR_ = 0.03), aMCI and SCD (z = -3.2, p_FDR_ = 0.003), aMCI and naMCI (z = -3.1, p_FDR_ = 0.003) (See Supplementary Figure 2).

Regional hypothalamic volume difference across different cognitive groups
*Anterior Superior Hypothalamus:*

Robust group differences were found bilaterally (left: χ² = 32.6, p < 0.001; right: χ² = 36.7, p < 0.001). The AD group exhibited significantly reduced volumes compared to MD-MCI (left: Z = -2.6, pFDR = 0.01; right: Z = -3.2, pFDR = 0.002), SD-MCI (left: Z = -4.6, pFDR < 0.001; right: Z = -4.2, pFDR < 0.001), and SCD (left: Z = -4.5, pFDR < 0.001; right: Z = -5.5, pFDR < 0.001) groups on both hemispheres. Additionally, the MD-MCI group showed smaller volumes than the SCD group bilaterally (left: Z = -3.3, pFDR = 0.001; right: Z = -4.0, pFDR < 0.001) and smaller left-sided volume compared with the SD-MCI group (left: Z = -3.5, pFDR = 0.001) (See Supplementary Figure 3).

*Anterior Inferior Hypothalamus:*

Significant bilateral group differences were also observed (left: χ² = 13.5, p = 0.004; right: χ² = 15.6, p = 0.001). The AD group had markedly smaller volumes than SD-MCI (left: Z = -3.0, pFDR = 0.009; right: Z = -3.4, pFDR = 0.004) and SCD (left: Z = -3.1, pFDR = 0.01; right: Z = -3.1, pFDR = 0.006) groups on both sides. On the right side, the MD-MCI groups exhibited a significant declined volume than SD-MCI group (Z = -2.5, pFDR = 0.03) (See Supplementary Figure 4).

*Tubular Superior Hypothalamus:*

Group differences were significant bilaterally (left: χ² = 15.1, p = 0.002; right: χ² = 22.4, p < 0.001). The AD group had significantly reduced bilateral volumes compared to the MD-MCI (left: Z = -2.2, pFDR = 0.04; right: Z = -3.1, pFDR = 0.003), SD-MCI (left: Z = -3.6, pFDR = 0.002; right: Z = -4.0, pFDR < 0.001), and SCD (left: Z = -3.0, pFDR = 0.008; right: Z = -4.4, pFDR < 0.001) groups. Besides, the MD-MCI group showed smaller volumes than SD-MCI group on left hemisphere (Z = -2.4, pFDR = 0.03) and SCD group on right hemisphere (Z = -2.3, pFDR = 0.04) (See Supplementary Figure 5).

*Tubular Inferior Hypothalamus:*

Though significant group difference was identified on left hemisphere (χ² = 8.7, p = 0.03), it did not pass the post-hoc pairwise comparison. No significant group difference was identified on right hemisphere (χ²= 6.7, p = 0.08) (See Supplementary Figure 6).

*Posterior Hypothalamus:*

Significant bilateral differences were found (left: χ² = 10.8, p = 0.01; right: χ² = 10.0, p = 0.02), with the AD group showing reduced volumes compared to MD-MCI (left: Z = -2.5, pFDR = 0.02; right: Z = -2.6, pFDR = 0.02), SD-MCI (left: Z= -2.7, pFDR = 0.02; right: Z = -2.8, pFDR = 0.02), and SCD (left: Z = -3.3, pFDR = 0.007; right: Z = -3.1, pFDR = 0.01) groups (See Supplementary Figure 7).

Correlations between hypothalamic volume and hippocampal & thalamic volume

Hippocampus: For hypothalamus subregions, anterior superior (rho = 0.5, pFDR < 0.001) and tubular superior (rho = 0.49, pFDR < 0.001) were mostly linked to hippocampus volume, followed by posterior (rho = 0.47, pFDR < 0.001), anterior inferior (rho = 0.44, pFDR < 0.001) and tubular inferior (rho = 0.4, pFDR < 0.001) (Supplementary Figure 8).

Thalamus: Among the hypothalamus subregions, anterior superior (rho = 0.52, pFDR < 0.001) exhibited the strongest associations with thalamic volume, followed by the tubular superior (rho = 0.51, pFDR < 0.001), posterior (rho = 0.46, pFDR < 0.001), anterior inferior (rho = 0.44, pFDR < 0.001) and tubular inferior (rho = 0.35, pFDR < 0.001) subregion (Supplementary Figure 9).

Hypothalamic Volume and PSQI

Sleep latency was associated with entire hypothalamus (left: rho = 0.10, pFDR = 0.019; right: rho = 0.15, pFDR < 0.001). For sleep efficiency, compared with individuals reported more than 85% sleep efficiency (β = 0.0545, 95% CI: (0.0539, 0.0551)), hypothalamus volume was larger in individuals with less than 65% sleep efficiency (β = 0.0565, 95% CI: (0.0554, 0.0577), p = 0.01) and those with 75%-84% sleep efficiency (β = 0.0559, 95% CI: (0.0549, 0.0569), p = 0.04).

There were small positive correlations between sleep latency and right entire hypothalamus volume (rho = 0.12, p_FDR_ = 0.005), and right tubular superior subregion volume (rho = 0.14, p_FDR_ < 0.001). There were small positive correlations between sleep duration and right entire hypothalamus volume (rho = 0.11, p_FDR_ = 0.01), and right tubular superior subregion volume (rho = 0.12, p_FDR_ = 0.006). There were small positive correlations between sleep efficiency and right entire hypothalamus volume (rho = 0.11, p_FDR_ = 0.01), and for hypothalamic subunits, right tubular superior (rho = 0.09, p_FDR_ = 0.0499) and inferior subregion volume (rho = 0.11, p_FDR_ = 0.01). There were small positive correlations between sleep disturbance right tubular superior (rho = 0.1, p_FDR_ = 0.03) and left posterior subregion volume (rho = 0.09, p_FDR_ = 0.03). There were no associations between hypothalamus volume and scores of sleep quality (Component 1), medication (Component 6) and daytime function (Component 7).

Across SCD, SD- and MD-MCI groups, small positive correlations between PSQI global score and the right hypothalamus volume (rho = 0.15, p_FDR_ < 0.001). Sleep latency was associated with entire hypothalamus (left: rho = 0.09, p_FDR_ = 0.033; right: rho = 0.14, p_FDR_ < 0.001). Sleep duration was positively related to whole hypothalamus volume (left: rho = 0.09, p_FDR_ = 0.043; right: rho = 0.13, p_FDR_ = 0.002). Sleep efficiency was correlated with right hypothalamus (rho = 0.14, p_FDR_ = 0.001) (See supplementary Figure 10).

Hypothalamus volume and individual PSG metrics

*Sleep efficiency:* There was no relationship between hypothalamus volume and sleep efficiency in the whole sample. However, within specific cognitive groups, negative correlation was found between whole left hypothalamus volume and sleep efficiency in the AD group (rho = -0.60, p_FDR_ = 0.04) (See Supplementary Figure 11,12).

Sleep latency: There was no relationship between hypothalamus volume and sleep latency in the whole sample. In the AD group, a positive association was found between sleep latency and whole left volume (rho = 0.60, p_FDR_ = 0.04), for subregions, sleep latency was related to volume of the tubular inferior hypothalamus (rho = 0.67, p_FDR_ = 0.02) (See Supplementary Figure 11,12).

REM latency: No association was found between hypothalamus volume.

*REM sleep:* No significant relationship was found between hypothalamic volume and sleep efficiency across the entire sample. In the SD-MCI group, a positive association was identified between REM sleep proportion and the volume of the entire left hypothalamus (rho = 0.40, p_FDR_ = 0.03), as well as the anterior superior region (rho = 0.54, p_FDR_ < 0.001) (See Supplementary Figure 11,12). 
 
*Light sleep:* There was no relationship between hypothalamus volume and light sleep proportion in the whole sample. In MD-MCI group, there were negative correlations between N1 sleep proportion and anterior inferior volume (left: rho = -0.31, p_FDR_ = 0.009) (See Supplementary Figure 11,12).  
 
*Slow wave sleep:* Across the entire sample, there was a significant positive association between both-side hypothalamus volume and N3 sleep proportion (left: rho = 0.17, p_FDR_ = 0.04; right: rho = 0.17, p_FDR_ = 0.04). There was also a weak correlation between left tubular inferior volume and N3 sleep proportion (rho= 0.17, p_FDR_ = 0.04). In SCD group, there were positive correlations between N3 sleep proportion and whole right hypothalamus volume (rho = 0.32, p_FDR_ = 0.049). In the AD group, a positive association was found between SWS proportion and posterior (rho = 0.61, p_FDR_ = 0.04) (See Supplementary Figure 11,12).

Hypothalamus volume and individual OSA metrics

Across the whole sample, there were negative correlations between SPO_2_ and whole hypothalamus volume (left: rho = -0.17, p_FDR_ = 0.03; right: rho = -0.19, p_FDR_ = 0.02). For subregions, SPO_2_ is related to the volume of anterior superior (rho = -0.19, p_FDR_ = 0.02), tubular part (superior: left: rho = -0.20, p_FDR_ = 0.01; inferior: left: rho = -0.18, p_FDR_ = 0.03) (See Supplementary Figure 13).

Correlations between hippocampus volume and PSG PCs
In the whole sample, no correlation was found between hippocampus volume and PCs.
PC1: (β = 0.95, SE = 1.19, t = 0.80, p_FDR_ = 0.54)
PC2: (β = -0.74, SE = 0.94, t = -0.79, p_FDR_ = 0.54)
PC3: (β = 1.70, SE = 0.81, t =2.11, p_FDR_ = 0.18) (ps: p = 0.0357 before multiple comparison)
PC4: (β = -1.09, SE = 0.73, t = -1.50, p_FDR_ = 0.33)
PC5: (β = -0.31, SE = 0.69, t = -0.45, p_FDR_ = 0.65)

Correlations between thalamus volume and PSG PCs

In the whole sample, no correlation was found between thalamus volume and PCs.
PC1: (β = -0.67, SE = 0.88, t = -0.76, p_FDR_ = 0.70)

PC2: (β = -0.53, SE = 0.69, t = -0.77, p_FDR_ = 0.70)
PC3: (β = 1.01, SE = 0.60, t = 1.69, p_FDR_ = 0.46)

PC4: (β = -0.31, SE = 0.54, t = -0.59, p_FDR_ = 0.70)
PC5: (β = 0.06, SE = 0.51, t = 0.12, p_FDR_ = 0.91)

Hypothalamus volume moderates the correlation between hippocampus volume and verbal memory performance

Anterior inferior: Volume of anterior inferior hypothalamus marginally moderated the correlation between hippocampal volume and verbal memory performance, with the interaction effect approaching statistical significance (β = -0.06, SE = 0.04, t = -1.9, p = 0.0573).

Tubular superior: Volume of tubular inferior hypothalamus marginally did not moderate the correlation between hippocampal volume and verbal memory performance (β = -0.05, SE = 0.04, t = -1.4, p = 0.151)

Tubular inferior: Volume of tubular inferior hypothalamus marginally moderated the correlation between hippocampal volume and verbal memory performance, with the interaction effect approaching statistical significance (β = -0.07, SE = 0.04, t = -1.9, p = 0.061).

Posterior: Volume of posterior hypothalamus marginally moderated the correlation between hippocampal volume and verbal memory performance, with the interaction effect approaching statistical significance (β = -0.07, SE = 0.03, t = -1.9, p = 0.055).
